# Amelioration of non-alcoholic fatty liver disease by targeting G protein-coupled receptor 110: A preclinical study

**DOI:** 10.1101/2022.12.07.22283206

**Authors:** Mengyao Wu, Tak-Ho Lo, Liping Li, Jia Sun, Chujun Deng, Ka-Ying Chan, Xiang Li, Steve Ting-Yuan Yeh, Jimmy Tsz Hang Lee, Pauline Po Yee Lui, Aimin Xu, Chi-Ming Wong

**Author notes:** Competing Interests: None. Abbreviations: ADGRE1, adhesion G Protein-Coupled Receptor E1; ADV, adenovirus; ALT, alanine aminotransferase; ASO, antisense oligonucleotide; AST, aspartate aminotransferase; cDNA, complementary DNA; CHO, cholesterol; DIO, diet-induced obesity; FGF21, fibroblast growth factor 21; GPCR, G protein coupled receptor; GPR110, G-protein coupled receptor 110; EWAT, epididymal white tissue; FFA, free fatty acids; GFP, green fluorescent protein; HCC, hepatocellular carcinoma; HDL, high-density lipoprotein; H&E, hematoxylin and eosin; HFD, high-fat diet; HOMA-IR, Homeostatic Model Assessment of Insulin Resistance; KEGG, Kyoto Encyclopedia of Genes and Genomes; LDL, low-density lipoprotein; MUFA, monounsaturated fatty acids; NAFLD, non-alcoholic fatty liver disease; NASH, non-alcoholic steatohepatitis; NCD, non-communicable diseases; NPC, Non-parenchymal cells; ORO, Oil Red O; SCD1, stearoyl-CoA desaturase-1; rAAV, recombinant adeno-associated virus; STC, standard chow diet; TG triglyceride.

## Abstract

**Background:** Recent research has shown that the G protein-coupled receptor 110 (GPR110) is an oncogene. The evidence mainly based on high expression of GPR110 in numerous cancer types; and knockdown GPR110 can reduced the cell migration, invasion, and proliferation. GPR110 is, however, mostly expressed in the liver of healthy individuals. The function of GPR110 in liver has not been revealed. Interestingly, expression level of hepatic GPR110 is dramatically decreased in obese subjects. Here, we examined whether GPR110 has a role in liver metabolism.

**Methods:** We used recombinant adeno-associated virus-mediated gene delivery system and antisense oligonucleotide to manipulate the hepatic GPR110 expression level in diet-induced obese mice to investigate the role of GPR110 in hepatic steatosis. The clinical relevance was examined using transcriptome profiling and archived biopsy specimens of liver tissues from non-alcoholic fatty liver disease (NAFLD) patients with different degree of fatty liver.

**Results:** The expression of GPR110 in the liver was directly correlated to fat content in the livers of both obese mice and NAFLD patients. Stearoyl-coA desaturase 1 (SCD1), a crucial enzyme in hepatic de novo lipogenesis, was identified as a downstream target of GPR110 by RNA-sequencing analysis. Treatment with the liver-specific SCD1 inhibitor MK8245 and specific shRNAs against SCD1 in primary hepatocytes improved the hepatic steatosis of GPR110-overexpressing mice and lipid profile of hepatocytes, respectively.

**Conclusions:** These results indicate GPR110 regulates hepatic lipid metabolism through controlling the expression of SCD1. Down-regulation of GPR110 expression can potentially serve as a protective mechanism to stop the over-accumulation of fat in the liver in obese subjects. Overall, our findings not only reveal a new mechanism regulation the progression of NALFD, but also proposed a novel therapeutic approach to combat NAFLD by targeting GPR110.

**Fundings:** This work was supported in part by National Natural Science Foundation of China 81870586 (CMW), 82270941 and 81974117 (JS), Area of Excellence AoE/M-707/18 (AX and CMW), and General Research Fund 15101520 (CMW).

**Highlights:**

- GPR110 regulates hepatic lipid metabolism.
- High level of hepatic GPR110 aggravates the progression of NAFLD by inducing SCD1 expression.
- Reduction in hepatic GPR110 is required to alleviate the progression of NAFLD.
- Targeting hepatic GPR110 improves hepatic steatosis.

## 1. Introduction

Liver is a vital organ as it is the site for undergoing a number of crucial physiological processes including digestion, metabolism, immunity and storage of nutrients [1]. Over-storage of lipid in the hepatocytes not caused by alcohol is known as non-alcoholic fatty liver disease (NAFLD), which is the most common liver pathological condition with a worldwide prevalence of 25% [2]. The development of NAFLD is contributed by many factors such as lipid metabolism disorders, over- or mal-nutrition, inflammation, virus infection, or liver injuries [3]. NAFLD usually does not entail any symptoms at early stages. However, if left untreated, NAFLD accounts for approximately 85% of all chronic noncommunicable diseases (NCDs), such as type 2 diabetes mellitus (T2DM), cardiovascular disease (CVD) and chronic kidney disease (CKD) [4, 5]. In addition, NAFLD may progress to non-alcoholic steatohepatitis (NASH) with fibrosis, cirrhosis or even hepatocellular carcinoma (HCC) [6]. Therefore, NAFLD imposes high economic and social burdens in terms of work productivity, health-related life quality and use of healthcare resources [5].

Improvement in managing NAFLD helps to resolve at least partially the progression of these diseases. Stopping the progression of NAFLD by lifestyle modifications such as increasing physical exercise activity and reduction of hypercaloric diet are only effective during the early stages before there is fibrosis. No medication is available to reverse the excessive fat storage in the liver once NASH developed. Therefore, it is urgently needed to unravel the mechanisms of NAFLD in order to accelerate the development, implementation, and explore new targets for the development of diagnostic tests and cost-effective therapies.

G protein-coupled receptors (GPCRs) are the largest and most diverse family of membrane receptor that play important roles in regulating most cellular and physiological processes [7]. GPCRs are major targets for currently approved drugs [8, 9]. A few GPCRs have been shown to play key roles in NAFLD and modulating their activities to ameliorate liver-related metabolic syndrome was proposed as NAFLD treatment [10, 11]. However, currently proposed targets for GPCR-medicated NAFLD treatment are not exclusively expressed in hepatocytes, thus the potential side effects on other organs should be considered.

Human GPR110 was identified by phylogenetic analysis based on highly conserved amino acid sequences of the G protein coupled receptor transmembrane domains in 2002 [12]. Mouse ortholog of hGPR110 was identified by the same research team two years later [13] and various splice variants were detected in deep sequencing experiments [14, 15]. So far, most GPR110 related studies focused on its tumorigenicity. In general, overexpression of GPR110 was observed in various cancers, and it was required to promote cancer cell survival, proliferation, and migration [16–21]. Therefore, it was suggested that targeting GPR110 may represent a new therapeutic strategy for anti-cancer treatment. It was also reported that GPR110 is required for proper fetal brain development and amelioration of neuroinflammation [22]. However, GPR110 is predominantly expressed in health adult livers. The function of hepatic GPR110 remains unexplored.

In this study, we provide the first evidence that GPR110 induces the expression of SCD1, which contributes to NAFLD. By HFD-induced NAFLD mouse model [23], we also show that the repression of hepatic GRP110 expression is a potential protective mechanism of preventing over accumulation of lipid in liver. Importantly, our findings not only reveal a new mechanism regulation the progression of NALFD, but also proposed a novel therapeutic approach to combat NAFLD by targeting GPR110.

## 2. Methods

### 2.1. Animals

All animal procedures were approved by the Animal Subjects Ethics Sub-Committee of the Hong Kong Polytechnic University and were conducted in accordance with the guidelines of the Centralized Animals Facilities. In general, eight-week-old C57BL/6J male mice were housed in pathogen-free conditions at controlled temperature with a 12-hour light-dark cycle and access to food and water *ad libitum*. The eight-week-old male mice were divided into two groups and fed with either standard chow diet (STC, 18.3% protein, 10.2% fat, 71.5% carbohydrates, Research Diet Inc., New Brunswick, NJ, USA) or high-fat diet (HFD, 20% protein, 45% fat, 35% carbohydrates, Research Diets Inc., New Brunswick, NJ, USA) for 8 weeks. The sample size was calculated based on previous findings, suggesting that group sizes of n=7-8 would be sufficient [24].

The recombinant adeno-associated virus vector rAAV2/8 transduction was conducted as described previously [25, 26]. Briefly, mice were tail vein injected with 3×10^11^ rAAV2/8 vector harboring either green fluorescent protein (GFP) or GPR110. For antisense oligonucleotide (ASO) delivery, validated ASOs against mouse GPR110 were provided by Ionis Pharmaceuticals and injected subcutaneously once a week at 5 mg/kg body weight to the mice. For SCD1 inhibitor delivery, MK-8245 (MedChemExpress, NJ, USA) was gavaged at 10 mg/kg BW once a week [27]. All measurements were carried out in a randomized order.

### 2.2. Primary hepatocyte isolation and adenovirus infection

Primary hepatocytes from different groups of mice were isolated using a two-step perfusion method as previously described [28, 29]. Briefly, type II collagenase was perfused to mice at a flow rate of 10 ml/min. Liver was collected and mesh in serum-free DMEM and hepatocytes were pelleted by centrifugation. For the supernatant containing nonparenchymal cells (NPCs), gradient solutions of Percoll were used for extraction. For adenovirus viral infection experiments, serum-starved cells were infected with adenoviruses carrying mouse GPR110 cDNA to overexpress GPR110. Similar adenoviral vectors encoding the green fluorescent protein (GFP) gene were used as controls.

### 2.3. Cell culture and Luciferase Reporter Assay

HEK293 cells were cultured in DMEM supplemented with 10% FBS, 100 U/ml penicillin and 100 μg/ml streptomycin at 37 °C and 5% CO2. HEK293 cells were seeded in 6-well plates and were transfected with pGL3-SCD1 promoter and adenoviral vector expressing either GPR110 (ADV-GPR110) or GFP (ADV-GFP) by using the transfection reagent (#E4981, Promega, WI, USA), following the manufacturer’s instruction. DHEA (C3270, APExBIO, TX, USA) was added into cells at the concentration of 100 μM and incubated at 37 °C for 24h. For the luciferase reporter assay, pRL-TK (Renilla luciferase) reporter plasmid was used as a transfection control. The luciferase assays were performed by using the Dual-Luciferase Reporter Assay System (#E1960, Promega, WI, USA).

### 2.4. Biochemical analysis

For glucose profile measurement, the blood glucose and insulin level were measured by collecting the blood samples from the tip of the tail using Accu-Chek® glucometer (Roche Diagnostics, Indiana, USA) as described [30]. For the glucose tolerance test (GTT), insulin tolerance test (ITT) and pyruvate tolerance test (PTT), mice were fasted overnight prior to intraperitoneal injection of glucose (1 g/kg body weight (BW) (Sigma, St. Louis, MO), 0.75 U/kg BW insulin (Novolin R, Novo Nordisk, Bagsvaerd, Denmark) or 1 g/kg BW pyruvate (Sigma, St. Louis, MO). Blood glucose levels were measured from the tip of tail vein at 15, 30, 60, 90 and 120 minutes after injection. For plasma and hepatic lipid level, serum levels of triglyceride (TG) and total cholesterol (CHO) were measured using commercial kit (Biosino, biotechnology and science INC, China) according to the manufacturer’s instructions. Hepatic lipids were extracted using Folch methodology and liver extract was dissolved in ethanol for TG and CHO measurement. Both serum and hepatic levels of free fatty acid (FFA) were measured using commercial kit (Solarbio, China) according to the manufacturer’s instructions. For liver function assay, the alanine aminotransferase (ALT) and aspartate aminotransferase (AST) levels were measured in serum using commercial kits (Stanbio, USA) according to the manufacturer’s instructions.

### 2.5. Histopathologic and Western blot analysis

Hematoxylin and eosin (H&E) and Oil Red O (ORO) staining were performed on paraffin-embedded and frozen liver sections, respectively. Detailed procedures of H&E and ORO were described previously [31, 32]. Representative histopathological images were acquired with a light microscope (Olympus, Tokyo, Japan). Western blot analysis was performed as previously described [25, 26, 28, 29]. Briefly, total protein was extracted from tissues and cultured cells with RIPA lysis buffer (65 mM Tris-HCl pH 7.5, 150 mM NaCl, 1 mM EDTA, 1% NP-40, 0.5% sodium deoxycholate and 0.1% SDS). Protein samples were then separated by gel electrophoresis and then transferred to PVDF membranes (IPVH00010, Merck Millipore, CA, USA). The expression of protein was detected by a ChemiDoc MP Imaging System (bio-Rad, Hercules, CA, USA). Primary antibodies used were shown in Table S1.

### 2.6. Quantitative real time PCR analysis

Total RNA was extracted with RNAiso Plus (#9109, TakaRa Bio Inc., Shiga, Japan) as previously described. RNA was reverse transcribed into cDNA with PrimeScript^TM^ RT reagent Kit (#RR037, TakaRa Bio Inc., Shiga, Japan). cDNA was then amplified with TB green Premix Ex Taq^TM^ II (Til Rnase H Plus) (##RR820A, TakaRa Bio Inc., Shiga, Japan). The real-time PCR was conducted with a LightCycler 96 qPCR System (Roche, Basel, Switzerland). The relative quantity of the targeted RNA was calculated through normalization to the quantity of the corresponding GAPDH mRNA level. Detailed primer sequences were listed below in Table S2.

### 2.7. Microarray and RNA sequencing

The liver of mice fed with either STC (n=6) or HFD (n=6) for 8 weeks were sent to Kompetenzzentrum Fluoreszente Bioanalytik (Germany) for gene expression analysis using Affymetrix Mouse Exon 1.0 ST Array. RNA was extracted from the liver of mice treated with rAAV-GPR110 and ASO-GPR110 using RNeasy Kits (QIAGEN, Hilden, Germany) according to the instructions. RNA concentration was quantified using NanoDrop™ 2000 Spectrophotometer (Thermo Scientific, Waltham, USA) and RNA quality was assessed using Agilent 2100 Bioanalyzer (Agilent, Santa Clara, USA). 10 μg of total RNA from liver with RNA integrity number (RIN) greater than 7 was used in RNA-seq. RNA-seq was performed by BGI and analyzed by Dr. Tom system (BGI, Shenzhen, China). A heat map was created based on log2 transformed counts from different samples. To be included in the heat map, genes were required to have at least 1000 counts, totaled over all samples, where and the standard deviation of the log2 had to exceed two.

### 2.8. Human samples

Liver biopsy specimens were collected from 9 biopsy-proven NAFLD patients [33]. Liver sections with H&E staining were subjected to histological evaluation of steatosis. Simple steatosis was defined by the presence of macrovascular steatosis affecting at least 5% of hepatocytes without inflammatory foci and evidence of hepatocellular injury in the form of hepatocyte ballooning [34]. Individuals with a heavy alcohol-drinking history (≥40 g/day for up to 2 weeks), drug-induced liver disease and hepatitis virus infection were excluded from the study. Clinical parameters of individuals were summarized in Table S1. The human study is approved by the Zhujiang Hospital, Southern Medical University, Guangzhou, China (Number: 2019-KY-097-01). Written informed consent was obtained from participants prior to their inclusion in the study.

### 2.9. Statistical analysis

All experiments were performed at least twice, and each experimental group included n ≥ 7 mice. Representative data were presented as mean ± standard error of mean (SEM). All statistical analysis was performed with the Graphpad Prism software (version 9.0, CA, USA). Statistical differences among two groups were performed using the unpaired Student t test or Mann-Whitney tests for the comparison of variables with or without normal distribution, respectively. The correlation between the two groups was assessed by non-parametric Spearman’s test. For multiple comparisons between three or more groups, one-way ANOVA with Bonferroni correction was conducted. In all statistical comparisons, p values <0.05 were accepted as significant.

## 3. Results

### 3.1. GPR110 mainly expresses in liver of mice and its expression is downregulated after HFD treatment

Firstly, we used microarray analysis to examine the change in expression levels of hepatic GPCRs in mice after HFD treatment (Table S3). In this screening, we found that GPR110 is mainly expressed in liver and its expression is dramatically decreased in the HFD-fed mice as compared to their STC-fed littermates. Remarkably, in agreement with previous studies [20, 35], GPR110 is mainly expressed in the liver of adult mice (Figure 1A-B). We also checked the GPR110 protein expression in those tissues by Western blot analysis. GRP110 proteins were mainly detected in liver and kidney samples (Figure 1B). Next, we used cell fractionation to identify the GPR110 expressing cells in liver (Figure 1C). The CD11b mRNAs were used as markers for non-parenchymal cells (NPC), and albumin mRNA for hepatocytes. Our cell fractionation clearly demonstrated that GPR110 mRNA is mainly expressed in hepatocytes (Figure 1C). This finding was further supported by Western blot analysis (Figure 1D). Remarkably, after HFD treatment for 8 weeks, the expression level of hepatic GPR110 declined to almost undetectable level as examined by qPCR analysis (Figure 1E). In contrast, the mRNA levels of two NAFLD related markers, FGF21 and F4/80 (also known as ADGRE1) were highly induced in the livers of HFD fed mice [36, 37]. Western blot analysis was performed to confirm the declined expression of GPR110 is also observed in protein level in the livers of HFD-fed mice (Figure 1F, upper panel). Interestingly, HFD-treatment did not affect the renal GRP110 expression (Figure 1F, lower panel). Collectively, the hepatic, but not renal, GPR110 level is tightly regulated by nutritional status.

**Figure 1:**
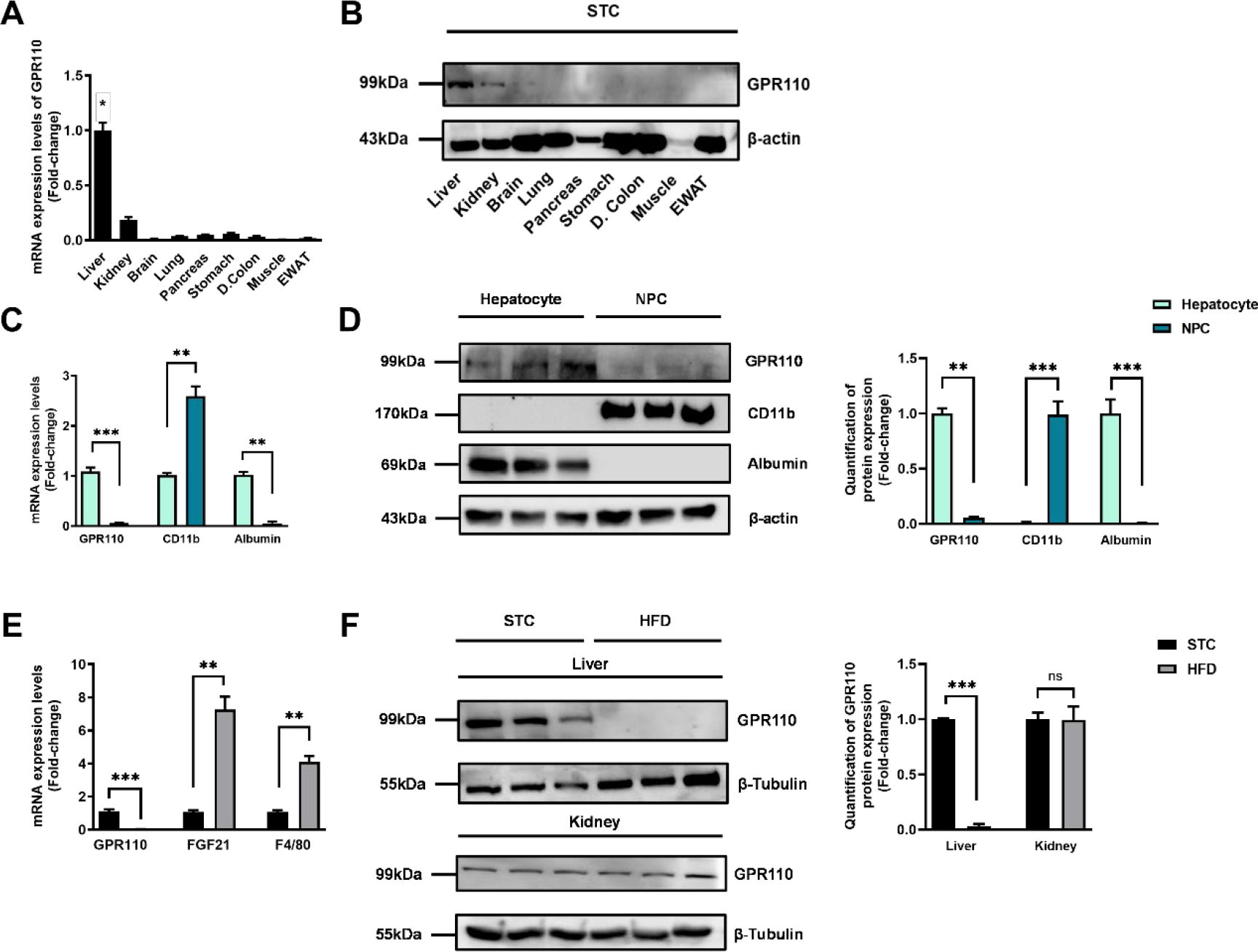
GPR110 is mainly expressed in the liver and its expression is downregulated after HFD treatment. Eight-week-old male C57BL/6J mice were fed with either STC or HFD for 8 weeks. (A) mRNA expression levels of GPR110 in different organs as determined by qPCR analysis (n = 5). (B) Representative immunoblotting analyses of GPR110 expression in different tissues of C57BL/J mice after STC for 8 weeks (n = 3). (C) mRNA expression levels of GPR110, CD11b and albumin in factions of hepatocyte or NPC isolated from STC-fed mice livers as determined by qPCR. (D) Left panel: representative immunoblotting analyses of GPR110, CD11b and albumin in fractions of hepatocytes or NPC isolated from mice livers fed with STC, each lane is a sample from different individual; right panel: quantification of protein expression levels of GPR110, CD11b and albumin. Protein expression levels were normalized to the expression of β-actin. The fraction of Hepatocytes was set as 1 for fold-change calculation. (E) mRNA expression levels of GPR110, FGF21 and F4/80 (served as HFD marker) in mice livers fed with either STC or HFD for 8 weeks as determined by qPCR. (F) Left panel: representative immunoblotting analyses of GPR110 in mice fed with either STC or HFD for 8 weeks; right panel: quantification of protein expression levels of GPR110. Protein expression levels were normalized to the expression of β-tubulin. The sample from STC mice were set as 1 for fold-change calculation. Each lane is a sample from different individual. GPR110, G-protein coupled receptor 110; STC, standard chow diet; HFD, high-fat diet; NPC, non-parenchymal cell. Data represents as mean ± SEM; n = 8 per group; repeated with three independent experiments; P value analyzed by two-tailed Student’s t test. *P < 0.05, **P < 0.01, ***P < 0.001.

### 3.2. Overexpression of GPR110 in hepatocytes accelerates metabolic dysregulation caused by HFD

Based on the dramatic difference in expression levels of hepatic GPR110 before and after HFD treatment, we hypothesized that downregulation of GPR110 in HFD-fed mice may be involved in the pathogenesis of fatty liver. To evaluate the impacts of high hepatic GPR110 level on liver metabolism, GPR110 was overexpressed in the hepatocytes of HFD-fed mice by liver-directed rAAV/ApoE- mediated gene expression system (Figure 2A and S1A). The overexpression of GPR110 in the livers of the mice were validated by qPCR (Figure 2B and S1B) and Western blot analysis (Figure S1C). We also confirmed that rAAV-mediated GPR110 overexpression was solely in hepatocytes, but not in NPC, by Western blot analysis after cell fractionation (Figure 2C). Renal GPR110 expression level was not affected by liver-directed rAAV/ApoE-mediated gene expression (Figure S1C).

**Figure 2:**
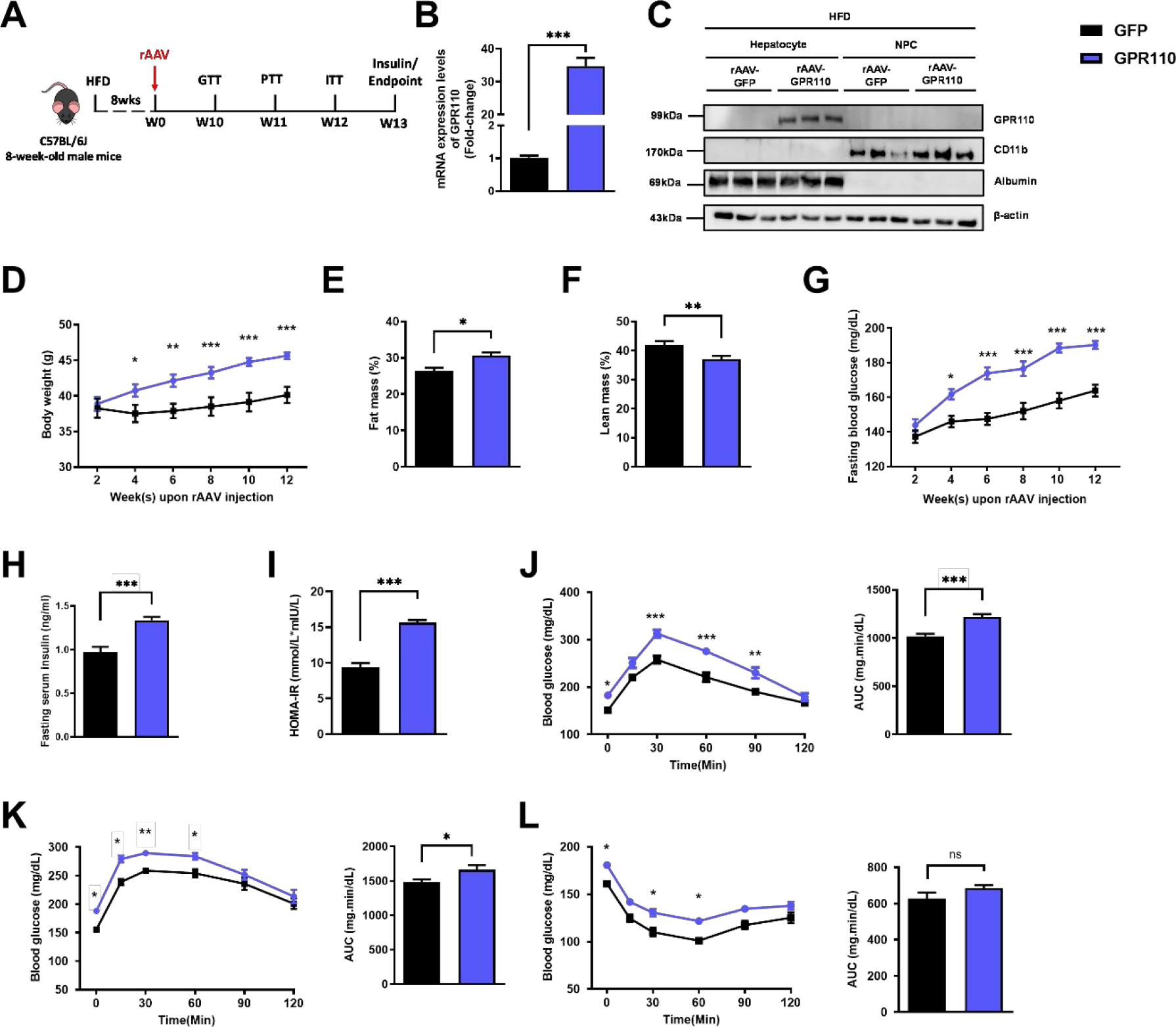
Overexpression of GPR110 in hepatocytes exaggerates metabolic dysregulation by HFD treatment. Eight-week-old male C57BL/6J mice were infected with 3×10^11^ copies of rAAV encoding GPR110 (rAAV-GPR110, i.v.) or control (rAAV-GFP, i.v.) and received HFD feeding, respectively. (A) Schematic illustration of viral treatments. (B) Hepatic mRNA expression levels of GPR110 from rAAV- GPR110 mice liver in fractions of hepatocyte or NPC isolated from HFD-fed mice livers as determined by qPCR. (C) Immunoblotting analyses of GPR110, CD11b and albumin from rAAV-GPR110 mice liver in factions of hepatocyte or NPC isolated from HFD-fed mice livers. Each lane is a sample from a different individual. n = 3 per group. (D) Body weight, (E) the percentage of fat mass and (F) lean mass were assessed in different groups. (G) Fasting blood glucose level were measured biweekly upon rAAV injection. (H) The fasting serum insulin level and (I) HOMA-IR index were measured and calculated according to the formula [Fasting blood glucose (mmol/l) × Fasting blood insulin (mIU/l)]/22.5 for the HFD-fed rAAV-GPR110 or rAAV-GFP mice at the end of the experiment (J) GTT (1 g/kg BW, left) and area under curve (AUC, right) of serum glucose at week 10. (K) PTT (1 g/kg BW, left) and AUC (right) of serum glucose at week 11. (L) ITT (0.5 U/kg BW, left) and AUC (right) of serum glucose at week 12. mRNA expression levels of the target genes were normalized to the expression of mouse GAPDH. rAAV-NC group was set as 1 for fold-change calculation. GPR110, G-protein coupled receptor 110; STC, standard chow diet; HFD, high-fat diet; NPC, non-parenchymal cell; BW, body weight; GTT, glucose tolerance test; PTT, pyruvate tolerance test; ITT, insulin tolerance test; AUC, area under curve; NC, negative control; HOMA-IR, homeostasis model assessment-estimated insulin resistance. Data represents as mean ± SEM; n = 8 mice per group; repeated with three independent experiments; P value analyzed by two-tailed Student’s t test. *P < 0.05, **P < 0.01, ***P < 0.001.

Overexpressing GPR110 in the liver of STC-fed mice did not affect body weight (Figure S1D), fasting glucose level (Figure S1E), fasting insulin level (Figure S1F) and homeostatic model assessment for insulin resistance (HOMA-IR; Figure S1G). There was only a slight increase at several time points in the glucose excursion curve in response to the GTT (Figure S1H) and hepatic glucose production induced by sodium pyruvate in PTT (Figure S1I). No change in insulin sensitivity was observed by insulin tolerance test between STC-fed rAAV-GFP and rAAV-GPR110 mice (Figure S1J).

However, under HFD treatment, rAAV-GPR110 mice gained more body weight (Figure 2D), and body fat mass (Figure 2E-F) than their rAAV-GFP controls. The HFD-fed rAAV-GPR110 mice also had higher fasting glucose level (Figure 2G), fasting insulin level (Figure 2H) and HOMA-IR (Figure 2I). Worsen glucose tolerance was observed in HFD-fed rAAV-GPR110 mice (Figure 2J). Overexpression of GPR110 in livers significantly increased hepatic glucose production induced in PTT (Figure 2K). ITT showed that the glucose levels in HFD-fed rAAV-GPR110 mice remained insensitive at 30 to 60 minutes after injection of insulin as compared to their control HFD-fed rAAV-GFP littermates (Figure 2L). In summary, we observed a mild impairment in glucose homeostasis associated with overexpressing GPR110 in the livers of STC-fed mice and more dramatical impairment was observed when the rAAV-GPR110 mice was challenged with HFD as compared to their rAAV-GFP controls.

3.3. Suppressing GPR110 improves glucose homeostasis in HFD-fed rAAV-GPR110 mice To confirm that the observations above were due to the rAAV-mediated overexpression of hepatic GPR110 in HFD-fed mice, we used two N-acetylgalactaosamine (GalNAc) conjugated antisense oligonucleotides (ASO-GPR110s) that bind to different regions of GPR110 mRNAs to knockdown the hepatic GPR110 expression in mice (Figure 3A and S2A). To avoid the observation is due to off-target effects, two different sequences of ASO were used. Chronic treatment of either ASO-GPR110s only lowered the hepatic, but not renal, GPR110 mRNA (Figure 3B and S2B) and protein (Figure S2C) levels. It is due to the fact that liver hepatocytes abundantly and specifically express the asialoglycoprotein receptor that binds and uptakes circulating glycosylated oligonucleotides via receptor-mediated endocytosis [38]. Knockdown hepatic GPR110 by ASO-GPR110s in STC-fed mice did not affect body weight (Figure S2D) and fasting glucose (Figure S2E), but slightly lowered insulin level (Figure S2F) and HOMR-IR (Figure S2G) as compared to their littermates injected with the negative control – scrambled antisense oligonucleotides (ASO-NC). No difference in the changes of glucose levels in GTT (Figure S2H), PTT (Figure S2I) and ITT (Figure S2J) for both ASO-GPR110s and ASO-NC groups under STC feeding conditions.

**Figure 3:**
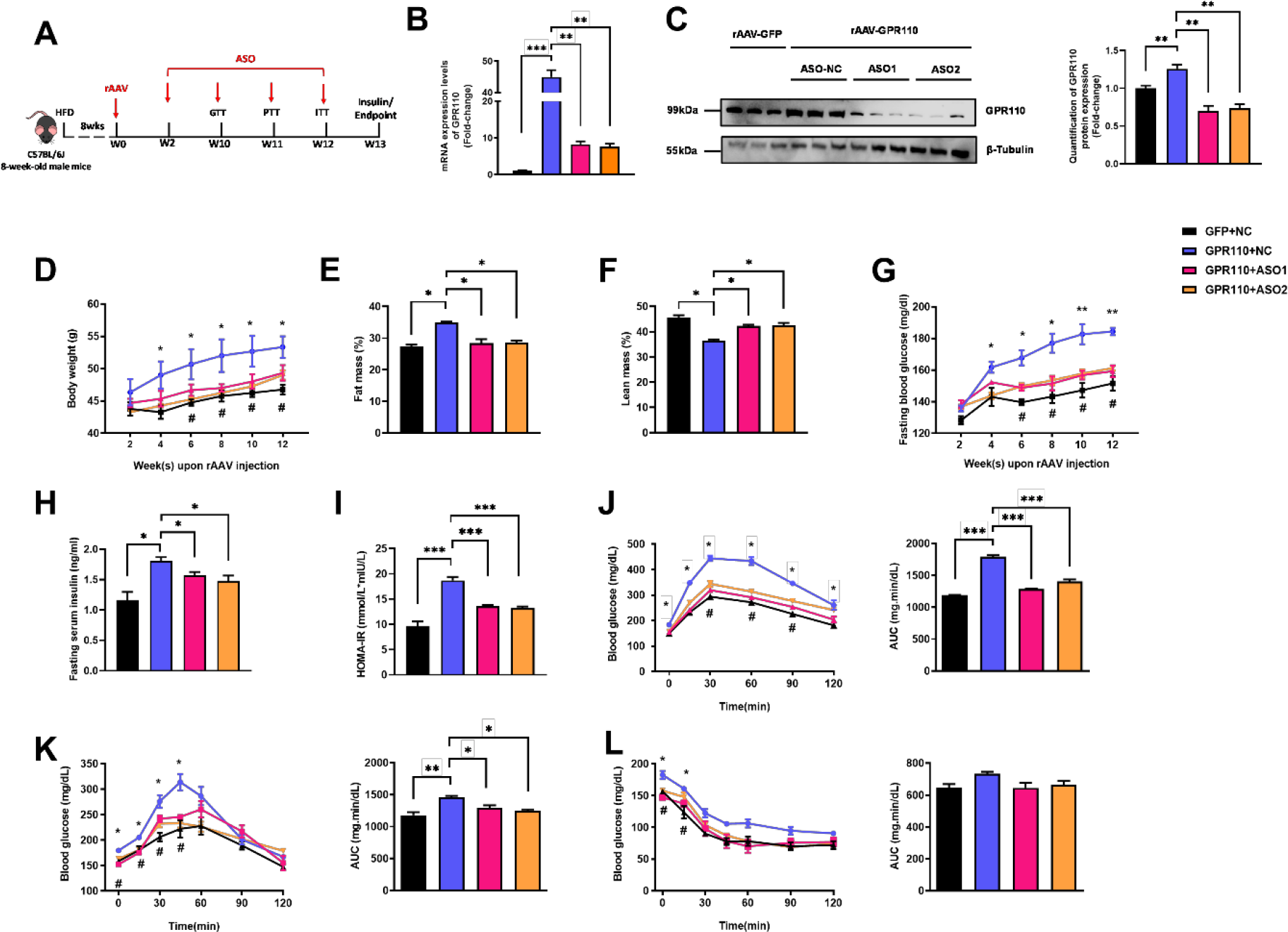
Deletion of hepatic GPR110 protects against diet-induced glucose intolerance in GPR110 overexpress mice. Eight-week-old male C57BL/6J mice were infected with either 3×10^11^ copies of rAAV encoding GPR110 (rAAV-GPR110, i.v.) or control (rAAV-GFP, i.v.) and two different sequences of GPR110 ASO (ASO1-GPR110, ASO2-GPR110, 5 mg/kg BW, one dose per week, s.c.) or scrambled control (ASO- NC, s.c.) received HFD feeding, respectively. (A) Schematic illustration of viral treatments. (B) Hepatic mRNA expression levels of GPR110 from different groups of mice received either GFP-NC, GPR110-NC, GPR110-ASO1 or GPR110-ASO2 fed with HFD, respectively, as determined by qPCR analysis. (C) Left panel: Immunoblotting analyses of GPR110 and β-tubulin from livers of HFD-fed rAAV-GFP or rAAV- GPR110 mice treated with either ASO-NC or ASO-GPR110. Each lane is a sample from a different individual. Right panel: quantification of protein expression levels of GPR110 and β-tubulin. Protein expression levels were normalized to the expression of β-tubulin. n = 3 per group. (D) Body weight was measured biweekly upon rAAV and ASO injection. (E) The percentage of fat mass and (F) the percentage of lean mass were measured at the end of the experiment. (G) The fasting blood glucose level of different groups were measured upon rAAV and ASO injection. (H) Fasting serum insulin level and (I) HOMA-IR index were measured and calculated according to the formula [Fasting blood glucose (mmol/l) × Fasting blood insulin (mIU/l)]/22.5 for the HFD-fed rAAV-GPR110 or rAAV-GFP mice at the end of the experiment. (J) GTT (1 g/kg BW, left) and AUC (right) of serum glucose at week 10. (K) PTT (1 g/kg BW, left) and AUC (right) of serum glucose at week 11. (L) ITT (0.5 U/kg BW, left) and AUC (right) of serum glucose at week 12. mRNA expression levels of the target genes were normalized to the expression of mouse GAPDH. rAAV-NC group was set as 1 for fold-change calculation. STC, standard chow diet; HFD, high-fat diet; ASO, antisense oligonucleotides; BW, body weight; GTT, glucose tolerance test; PTT, pyruvate tolerance test; ITT, insulin tolerance test; AUC, area under curve; NC, negative control; HOMA-IR, homeostasis model assessment-estimated insulin resistance. Data represents as mean ± SEM; n = 8 mice per group; repeated with three independent experiments; P value analyzed by two-tailed Student’s t test. *P < 0.05, **P < 0.01, ***P < 0.001.

In contrast, chronic ASO-GPR110 treatment for 4 weeks significantly decreased their body weight (Figure 3D), fat mass ratio (Figure 3E-F) fasting glucose level (Figure 3G), fasting insulin level (Figure 3H) and HOMA-IR (Figure 3I) in HFD-fed rAAV-GPR110 mice. In addition, treatment of ASO-GPR110s improved glucose tolerance, pyruvate tolerance and insulin sensitivity in HFD-fed rAAV-GPR110 mice as demonstrated by GTT (Figure 3J), PTT (Figure 3K) and ITT (Figure 3L) as compared to ASO-NC controls. In consistent to overexpressing GPR110 in livers, the depletion of hepatic GPR110 by ASOs improves glucose homeostasis in HFD-fed mice.

### 3.4. Treatment of ASO-GPR110s alleviates lipid abundance and liver damage in HFD-fed rAAV- GPR110 mice

We also checked the circulating lipid profiles of the mice. HFD-fed rAAV-GPR110 mice had higher circulating cholesterol (CHO; Figure 4A) and triglyceride (TG; Figure 4B) levels than HFD-fed rAAV- GFP littermates, but their circulating free fatty acid (FFA) levels were similar (Figure 4C). High-density lipoprotein (HDL) cholesterol level was decreased, and low-density lipoprotein (LDL) cholesterol level was increased in HFD-fed rAAV-GRP110-NC mice as compared to chronic ASO-GPR110 treatment group (Figure 4D).

**Figure 4:**
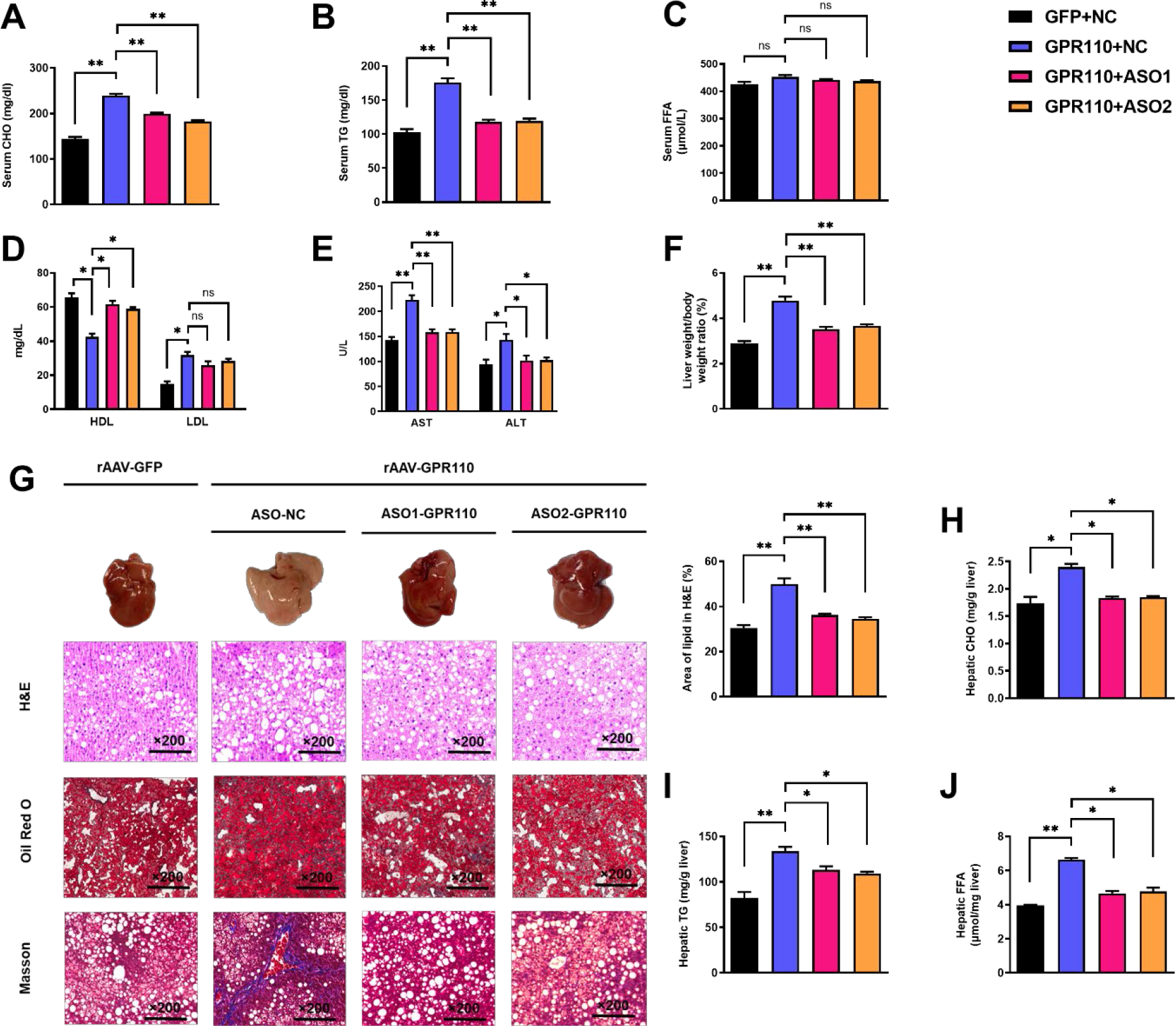
Up-regulation of hepatic GPR110 exaggerates liver steatosis in HFD mice fed with HFD while down-regulation of hepatic GPR110 protects mice from diet-induced liver lipid accumulation. Eight-week-old male C57BL/6J mice were infected with either 3×10^11^ copies of rAAV encoding GPR110 (rAAV-GPR110, i.v.) or control (rAAV-NC, i.v.) and two different sequences of GPR110 antisense oligonucleotides (ASO1-GPR110, ASO2-GPR110, 5 mg/kg BW, one dose per week, s.c.) or scrambled control (ASO-NC, s.c.) received HFD feeding, respectively. (A) Serum cholesterol (CHO), (B) serum triglyceride (TG) and (C) serum free fatty acid (FFA) levels were measured at week 13. (D) Serum high- density lipoprotein (HDL) and low-density lipoprotein (LDL). (E) The levels of serum aspartate transaminase (AST) alanine transaminase (ALT). (F) The ratio of the liver weight against body weight was calculated after sacrificing the mice from four different groups. (G) Representative gross pictures of liver tissues (upper panels), representative images of H&E (middle panels) and Oil Red O (lower panels) staining of liver sections (200 µm). The percentage of lipid area according to H&E staining (right panel); n = 3 per group. (H) Hepatic CHO, (I) hepatic TG and (J) hepatic FFA were normalized by the weight of liver samples used for lipid extraction. i.v., intravenous injection; s.c., subcutaneous injection; STC, standard chow diet; HFD, high-fat diet; ASO, antisense oligonucleotides; BW, body weight; CHO, cholesterol; TG, triglyceride; FFA, free fatty acid; HDL, high-density lipoprotein; LDL, low-density lipoprotein; AST, aspartate transaminase; ALT, alanine transaminase; H&E, hematoxylin-eosin. Data represents as mean ± SEM; n = 8 mice per group; repeated with three independent experiments; P value analyzed by two-tailed Student’s t test. *P < 0.05, **P < 0.01, ***P < 0.001.

Remarkably, chronic ASO-GPR110 treatment could lower circulating levels of the liver enzymes aspartate transaminase (AST) and alanine aminotransaminase (ALT), as markers of liver damage and hepatoxicity, in HFD-fed rAAV-GRP110-NC mice (Figure 4E). After sacrificing the mice, we examined their hepatic lipid profiles. We found the livers of HFD-fed rAAV-GRP110 mice were significantly heavier (Figure 4F) and paler (Figure 4G, upper panels) than the livers of their rAAV-GFP littermates and ASO- GPR110 treated rAAV-GRP110 mice. Consistent with these observations, HFD-induced lipid accumulation within hepatocytes were substantially more abundant in the livers of HFD-fed rAAV- GPR110 mice than the rAAV-GFP littermates as determined by haematoxylin and eosin (H&E) staining and Oil Red O staining (Figure 4G, upper and middle rows). Moreover, based on the Masson trichrome staining, more fibre extension and larger fibrous septa formation was observed for the liver samples of rAAV-GPR110 mice as compared to the livers from rAAV-GFP littermates (Figure 4G, lower row). These alterations were remarkably reduced after ASO-GPR110 treatments (Figure 4G). Like the circulating lipid profiles mentioned above, treatment of ASO-GPR110s for 8 weeks could improve the hepatic lipid profiles of rAAV-GRP110 mice in terms of CHO (Figure 4H), TG (Figure 4I) and FFA (Figure 4J). Altogether, overexpression of hepatic GPR110 in mice is sufficient to perturb lipid metabolism and hence the progression of NAFLD especially in obese subjects.

### 3.5. The metabolic dysregulation of rAAV-GRP110 mice is correlated to the upregulation SCD1 expression

To reveal the molecular mechanism underlying the involvement of hepatic GPR110 in NAFLD development, RNA-sequencing analysis was performed on RNA samples extracted from the livers of HFD-fed ASO-NC treated rAAV-GPF, ASO-NC treated rAAV-GPR110 and ASO-GRP110 treated rAAV-GPP110 mice. In the search for the molecular processes for metabolisms, several lipid metabolism-related genes were altered (Figure 5A-B). We subsequently used RT-qPCR to confirm the RNA sequencing results (Figure 5C). Among them, we are particularly interested in stearoyl CoA desaturase 1 (SCD1). SCD1 is a key lipogenic enzyme responsible for the rate-limiting step in the synthesis of monounsaturated fatty acids (MUFAs), such as oleate and palmitoleate, by forming double bonds in saturated fatty acids [39]. MUFAs serve as substrates for the synthesis of various kinds of lipids and increases in SCD1 activity is involved in the development of NAFLD, hypertriglyceridemia, atherosclerosis, and diabetes [40, 41].

**Figure 5:**
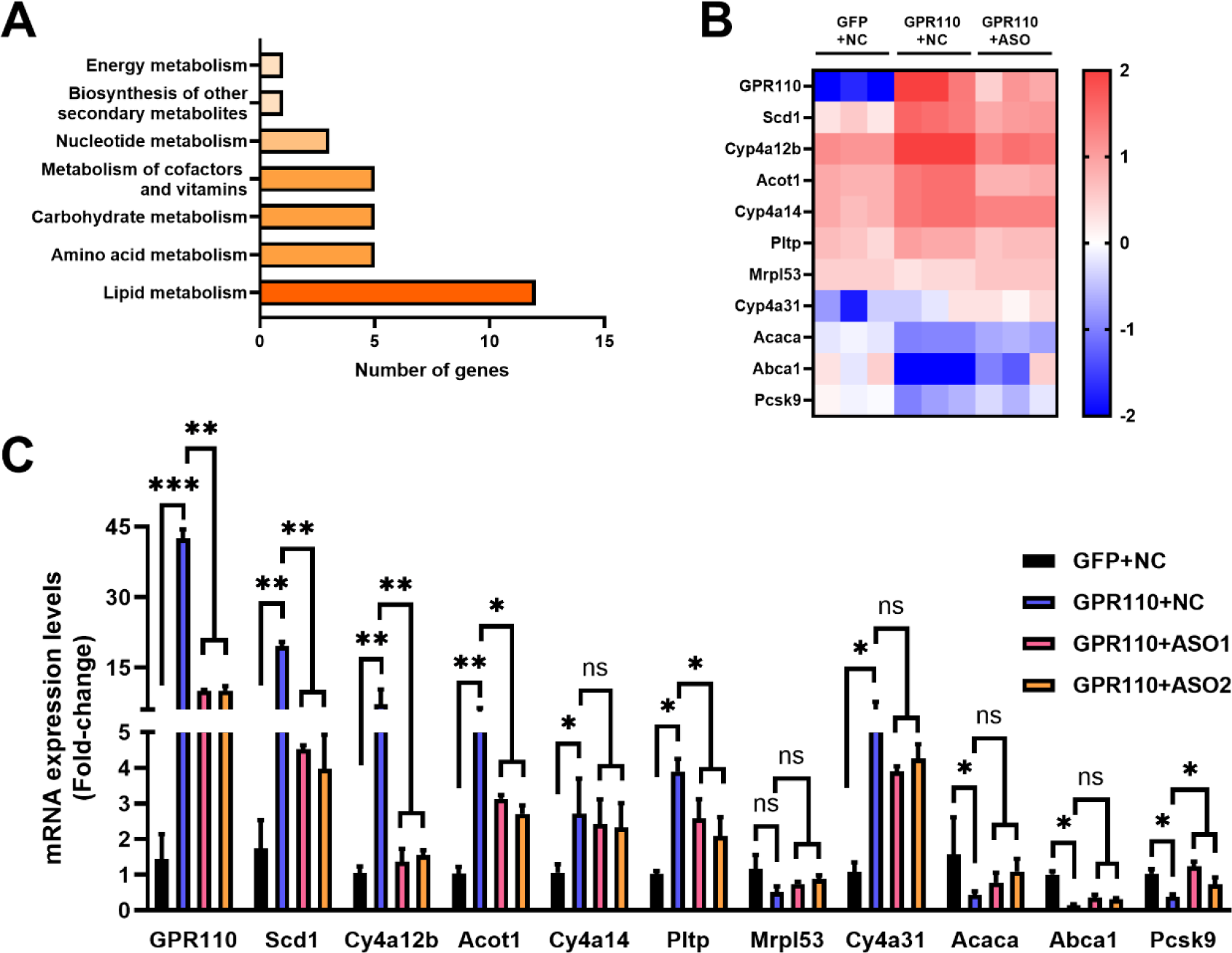
GPR110 is a major regulator of hepatic lipid metabolism. Eight-week-old male C57BL/6J mice were infected with either 3×10^11^ copies of rAAV encoding GPR110 (rAAV-GPR110, i.v.) or control (rAAV-NC, i.v.) and two GPR110 antisense oligonucleotides (ASO1-GPR110, ASO2-GPR110, 5 mg/kg BW, one dose per week, s.c.) or scrambled control (ASO-NC, s.c.) and received HFD feeding, respectively. Mice were sacrificed and mRNA of liver from each group were extracted and RNA-seq analysis was conducted. (A) KEGG pathway assay of differential mRNA transcripts in rAAV and ASO groups identified by RNA-seq. (B) Heat map show the Log2 scale fold change in the expression levels of a set of genes involved in lipid metabolism from RNA seq data of livers in HFD-fed mice treated by rAAV-GPR110 or rAAV-GPR110 plus GPR110-ASO1. n = 3 per group. (C) mRNA expression levels of genes according to the heatmap from different groups of mice received either GFP-NC, GPR110-NC, GPR110-ASO1 or GPR110-ASO2 fed with HFD, respectively, as determined by qPCR analysis, n = 6 mice per group. STC, standard chow diet; HFD, high-fat diet; i.v., intravenous injection; s.c., subcutaneous injection; ASO, antisense oligonucleotides; KEGG, Kyoto Encyclopedia of Genes and Genomes; GEO, gene expression omnibus; NAFLD, non-alcoholic fatty liver disease. Data represents as mean ± SEM; P value analyzed by two-tailed Student’s t test. *P < 0.05, **P < 0.01, ***P < 0.001.

### 3.6. The upregulation of SCD1 expression in liver is driven by the presence of GPR110

To confirm SCD1 expression is induced by GPR110, *in vitro* assays were performed by using adenovirus-mediated GPR110 expression system (ADV-GPR110) to overexpress GPR110 in primary hepatocytes isolated from STC-fed mice. After infection, the expressions of SCD1 mRNAs (Figure 6A) and protein (Figure 6B) were dramatically induced, but not in control group which was treated with ADV- GFP. In addition, the GPR110 specific ASOs can not only knockdown the GPR110 expression, but also induced the SCD1 expression by increasing GPR110 in the ADV-GPR110 primary hepatocytes (Figure 6A-B). *In vitro* luciferase reporter assay was performed to further validate the expression of SCD1 is transcriptional regulated by GPR110. We constructed plasmid harbouring luciferase gene driven by the mouse SCD1 promoter (-2000 to +100) and transfected into HEK293 cells. There was no change of luciferase activity of pGL3- SCD1 promoter-luciferase transfected HEK293 cells under the treatment of GPR110 ligand DHEA, unless the HEK293 cells were pre-infected with adenovirus overexpressing GPR110 (ADV-GPR110; Figure 6C). The overexpression of GPR110 in ADV-GPR110 infected cells and inductions of SCD1 mRNA expression by treatment of DHEA were also validated by qPCR (Figure 6D). The changes in hepatocyte lipid profiles by the expression level of GPR110 and SCD1 were also checked. In agreement with our *in vivo* studies’ findings, the overexpression of GPR110 increased the intracellular CHO (Figure 6E), TG (Figure 6F) and FFA (Figure 6G). Their increases could be completely repressed by ASO against GPR110 (Figure 6E-G) and partially repressed by overexpressing SCD1 specific shRNAs (Figure 6F-G). In summary, the transcription level of SCD1 is regulated by GPR110. GPR110 enhances the lipid accumulation by inducing SCD1 expression.

**Figure 6:**
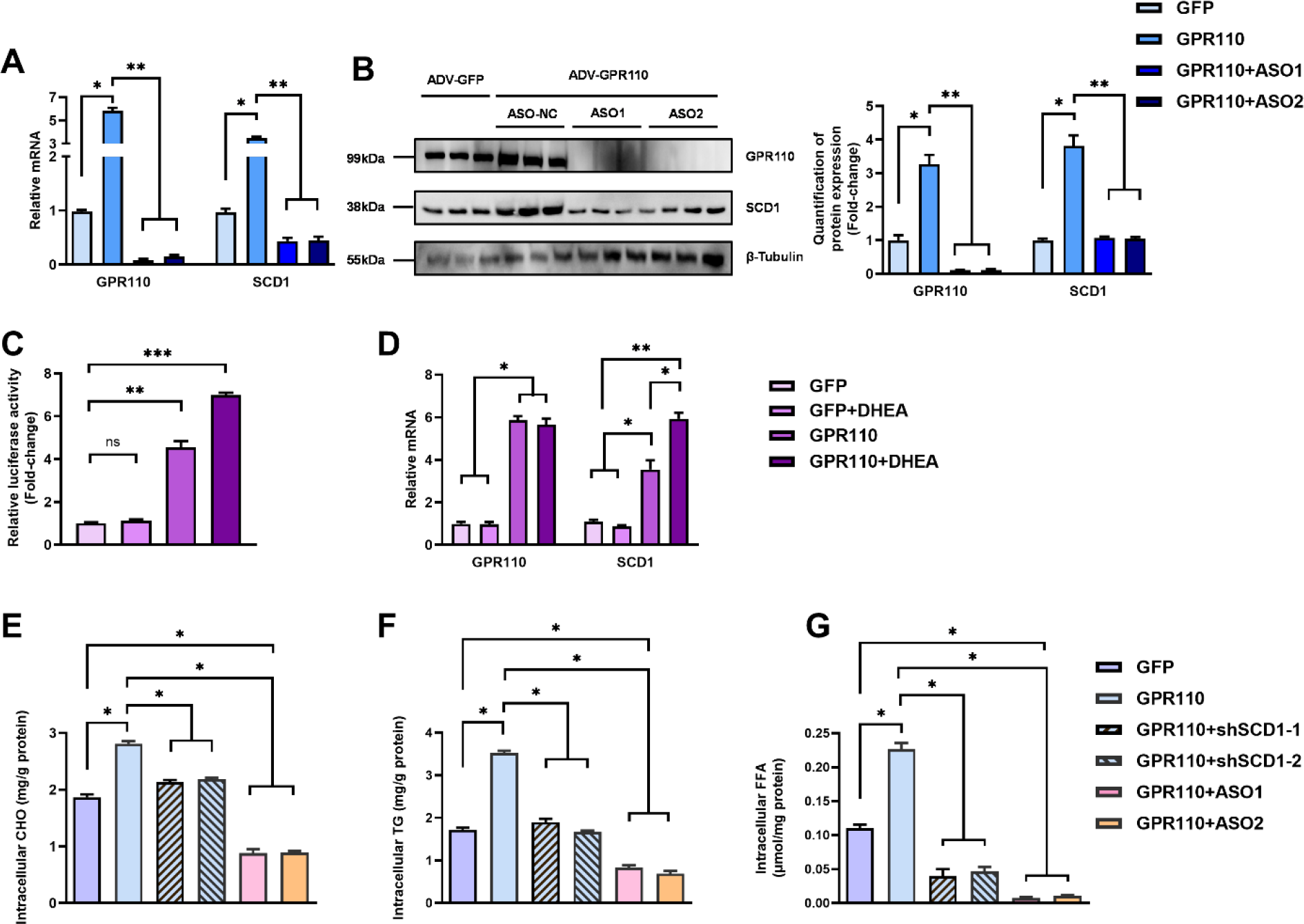
SCD1 expression is regulated by GPR110 in primary hepatocytes. Primary hepatocytes were isolated from eight-week-old male C57BL/6J mice with STC. (A) Primary hepatocytes were infected with either adenoviral vector expressing GPR110 (ADV-GPR110) or control adenovirus expressing GFP (ADV- GFP) 24h after plating, followed by transfection with ASO1-GPR110, ASO2-GPR110 or ASO-NC for another 6 hours. mRNA expression levels of GPR110 and SCD1 from different groups were assessed, as determined by qPCR analysis. (B) Left panel: immunoblotting analysis for the expression level of GPR110 and SCD1 from different groups of primary hepatocytes. Right panel: quantification of protein expression levels of GPR110 and SCD1. Protein expression levels were normalized to the expression of β-tubulin. Each lane is a sample from a different plate. Right panel: quantification of protein expression levels of GPR110, SCD1 and β-tubulin. n = 3 per group. Protein expression levels were normalized to the expression of β-tubulin. The samples for GFP were set as 1 for fold-change calculation. (C-D) HEK293 cells were infected with pGL3-SCD1 promoter-luciferase plasmid and adenoviral vector expressing GPR110 (ADV- GPR110) or GFP (ADV-GFP) for 48 h and DHEA was added to the transfected cells at the concentration of 100 μM for 24 h. Cell lysates were used for (C) luciferase assay or (D) qPCR analysis. Lysates from the cell co-transfection with pGL3-SCD1 promoter-luciferase plasmid and ADV-GFP without treatment of DHEA was set as 1 for fold-change calculation. (E-G) Primary hepatocytes were infected with either adenoviral vector expressing GPR110 (ADV-GPR110) or control ADV-GFP, followed by transfecting with scramble or shSCD1-1 or shSCD1-2 plasmids for another 72 h. Intracellular lipids were extracted and (E) CHO, (F) TG, and (G) FFA were assessed. STC, standard chow diet; i.v., intravenous injection; s.c., subcutaneous injection; ASO, antisense oligonucleotides. CHO, cholesterol; TG, triglyceride; FFA, free fatty acid. Data represents as mean ± SEM; n = 3 per group; repeated with three independent experiments; P value analysed by two-tailed Student’s t test. *P < 0.05, **P < 0.01, ***P < 0.001.

### 3.7. Inhibition of SCD1 in rAAV-GPR110 mice partially attenuates most metabolic dysregulations especially lipid profiles

To examine whether the up-regulation of hepatic SCD1 was the cause of metabolic dysregulation in rAAV-GRP110 mice, a liver-specific SCD1 inhibitor MK8245 were used to alleviate the metabolic dysregulation by overexpressing GPR110 in HFD-fed rAAV-GRP110 mice (Figure 7A) [27]. Chronic treatment of MK8245 for 11 weeks did not affect the expression of GPR110 mRNA (Figure 7B) and protein (Figure 7C) levels in rAAV-GRP110 mice. In agreement with previous studies showing that the chronic treatment of this SCD1 inhibitor improves various metabolic parameters including lipid and glucose profiles in various animal models [42], treatment of MK8245 lowered the body weight (Figure 7D), improved glucose homeostasis in term of fasting glucose level (Figure 7E) and HOMR-IR (Figure 7G), and performance in GTT (Figure 7H) and PTT (Figure 7I) of HFD-fed rAAV-GRP110 mice as compared to untreated littermates. But there was no change in insulin sensitivity as demonstrated by ITT (Figure 7J).

**Figure 7:**
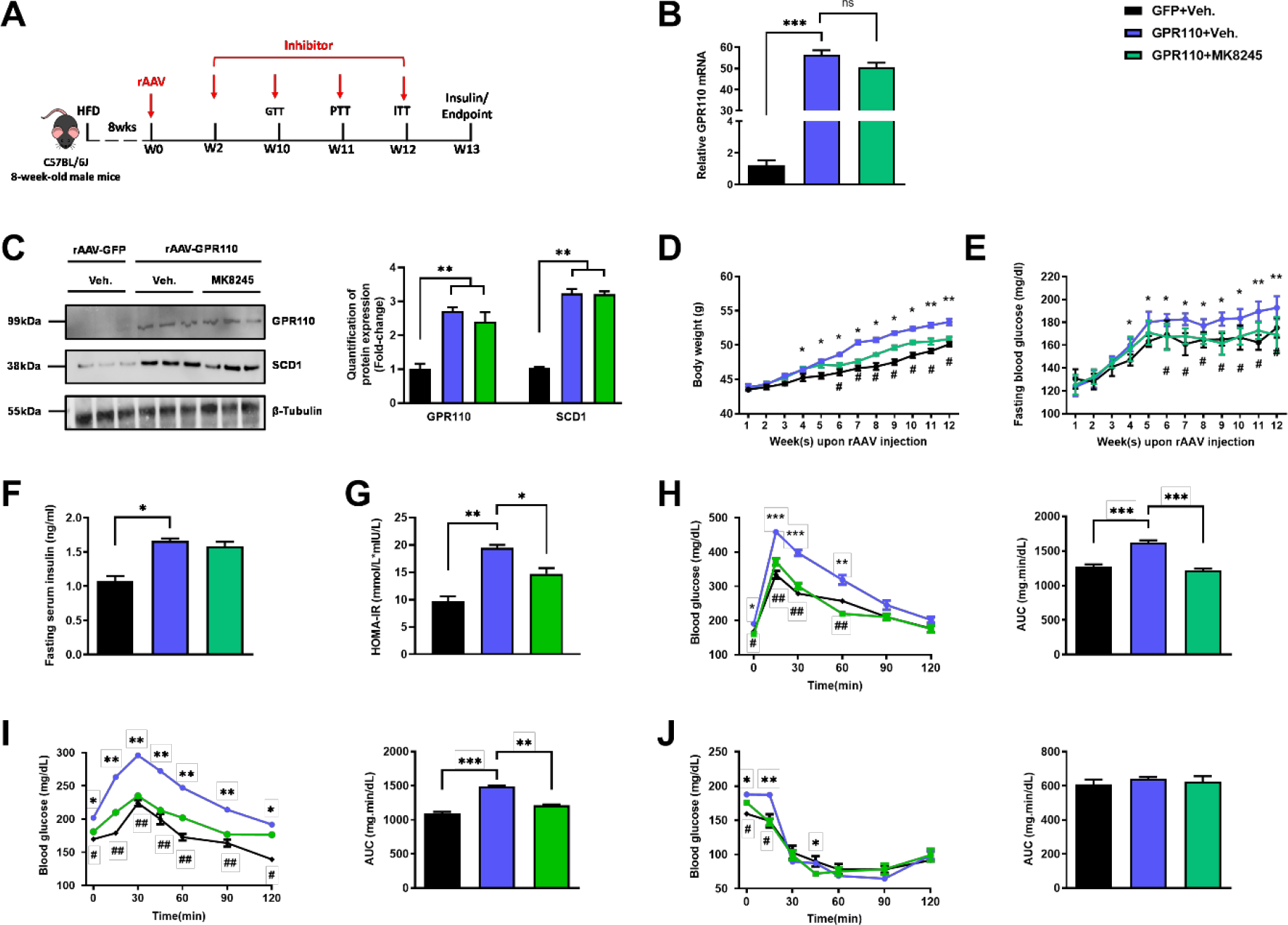
Inhibition of SCD1 alleviates the glucose impairment in mice with hepatic GPR110 overexpression. Eight-week-old male C57BL/6J mice were infected with either 3×10^11^ copies of rAAV encoding GPR110 (rAAV-GPR110, i.v.) or control (rAAV-GFP, i.v.) and SCD1 inhibitor (MK8245, 10 mg/kg BW/week, p.o.) or inhibitor vehicle (inhibitor-Veh., p.o.) received HFD feeding. (A) Schematic illustration of viral treatments. (B) Hepatic mRNA expression levels of GPR110 from different groups of mice received rAAV and inhibitor fed with HFD respectively, as determined by qPCR analysis. (C) Left panel: immunoblotting analysis for the hepatic protein expression level of GPR110 and SCD1 from different groups of mice fed with HFD. Right panel: quantification of protein expression levels of GPR110 and SCD1. Protein expression levels were normalized to the expression of β-tubulin. Each lane is a sample from a different individual; n = 3 per group. (D) Body weight and (E) fasting blood glucose level were measured at different weeks upon rAAV and inhibitor injection. (F) The fasting blood insulin level and (G) HOMA-IR index were measured and calculated according to the formula [Fasting blood glucose (mmol/l) × Fasting blood insulin (mIU/l)]/22.5 for the HFD-fed rAAV-GPR110 or rAAV-GFP mice at the end of the experiment. (H) GTT (1 g/kg BW, left) and AUC (right) of serum glucose at the week of 10. (I) PTT (1 g/kg BW, left) and AUC (right) of serum glucose at week 11. (J) ITT (0.5 U/kg BW, left) and AUC (right) of serum glucose at week of 12. mRNA expression levels of the target genes were normalized to the expression of mouse GAPDH. rAAV- NC group was set as 1 for fold-change calculation. HFD, high-fat diet; i.v., intravenous injection; p.o., oral administration; ASO, antisense oligonucleotides; BW, body weight; GTT, glucose tolerance test; PTT, pyruvate tolerance test; ITT, insulin tolerance test; AUC, area under curve; NC, negative control; HOMA- IR, homeostasis model assessment-estimated insulin resistance. Data represents as mean ± SEM; n = 8 mice per group; repeated with three independent experiments; P value analysed by two-tailed Student’s t test. *P < 0.05, **P < 0.01, ***P < 0.001.

MK8245 treatment also lowered the circulating CHO (Figure 8A) and TG (Figure 8B) levels almost to the levels of HFD-fed rAAV-GFP mice, but there was no change in circulating FFA level (Figure 8C). A relatively higher HDL can be found in the MK8245 group but there were no differences detected regarding the LDL level (Figure 8D). The AST and ALT levels were also alleviated in the MK8245 group compared to the rAAV-GPR110 littermates (Figure 8E). MK8245 treatment partially reduced the liver weight (Figure 8F), degree of paleness, severity of fibrosis (Figure 8G) and lipid accumulations (Figure 8H-J). To conclude, treatment of MK8245 could improve the lipid profiles and alleviate metabolic dysregulation caused by overexpression of hepatic GPR110 in mice.

**Figure 8:**
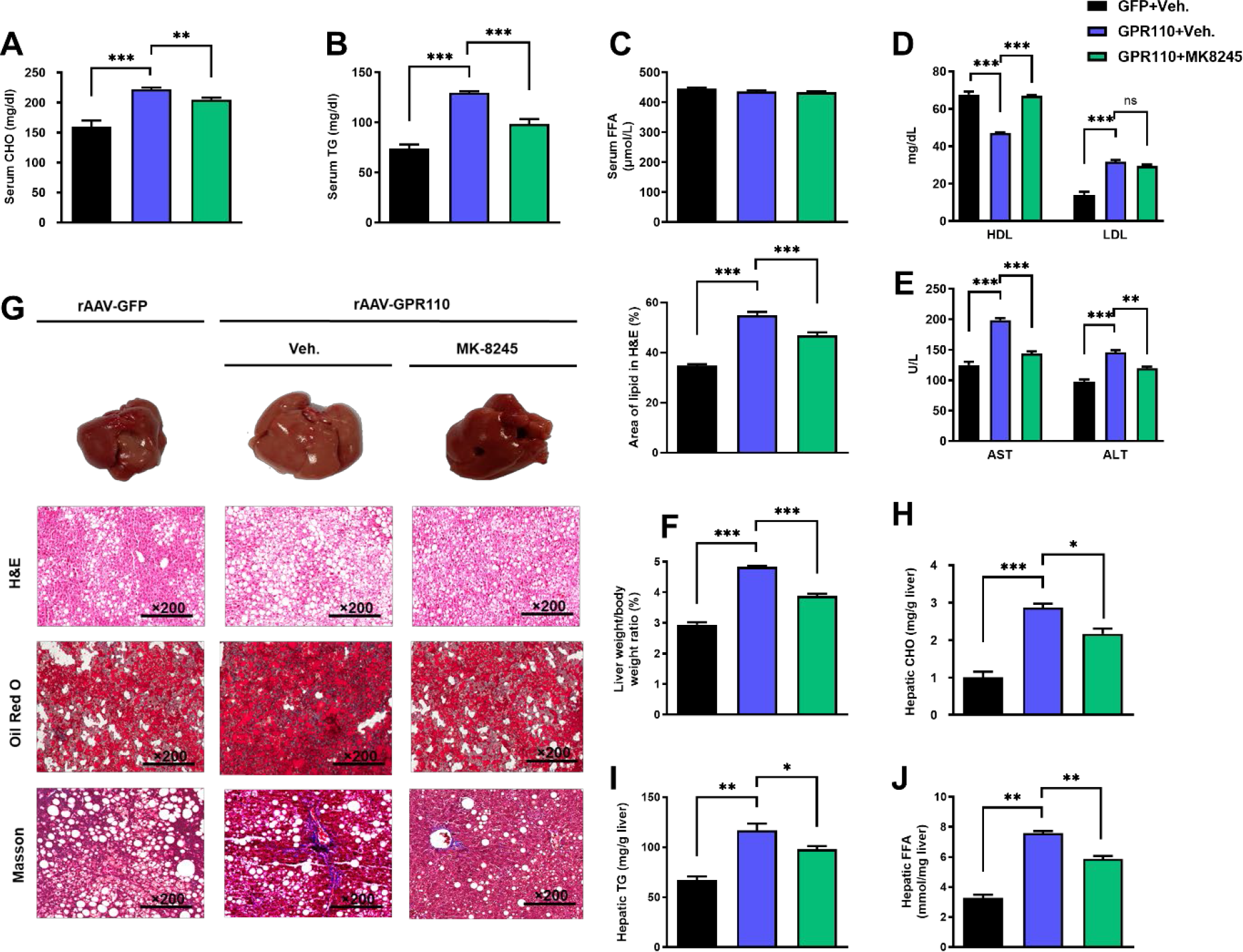
Inhibition of hepatic SCD1 partially alleviates the severity of hepatic steatosis in GPR110 overexpression mice. Eight-week-old male C57BL/6N mice were infected with either 3×10^11^ copies of rAAV encoding GPR110 (rAAV-GPR110, i.v.) or control (rAAV-GFP, i.v.) and administered with SCD1 inhibitor (MK8245, 10 mg/kg BW, p.o.) or inhibitor vehicle (inhibitor-Veh., p.o.) received HFD feeding. (A) Serum CHO, (B) serum TG and (C) serum FFA levels were measured at the end of experiment. (D) Serum HDL and LDL, (E) AST and ALT level of each group of mice were measured at the end of the experiment. (F) The ratio of the liver weight against body weight was calculated after sacrificing the mice from four different groups. (G) Representative gross pictures of liver tissues (upper panels), representative images of H&E (middle panels) and Oil Red O (lower panels) staining of liver sections (200 µm). The percentage of lipid area according to H&E staining (right panel); n = 3 per group. (H) Hepatic CHO, (I) hepatic TG and (J) hepatic FFA were normalized by the weight of liver samples used for lipid extraction. STC, standard chow diet; HFD, high-fat diet; i.v., intravenous injection; p.o., oral administration; CHO, cholesterol; TG, triglyceride; FFA, free fatty acid; HDL, high-density lipoprotein; LDL, low-density lipoprotein; AST, aspartate transaminase; ALT, alanine transaminase; H&E, hematoxylin-eosin. Data represents as mean ± SEM; n = 8 mice per group; repeated with three independent experiments; P value analyzed by two-tailed Student’s t test. *P < 0.05, **P < 0.01, ***P < 0.001.

### 3.8. Expression of GPR110 in liver is closely associated with hepatic steatosis in NAFLD patients

To evaluate the clinical relevance of our findings in mice, we first checked the expression level of GPR110 in human liver from a publicized transcriptome dataset Gene Expression Omnibus (GEO; Profile # GDS4881) with human liver biopsy of different phases from control to NAFLD [43]. Healthy obese subjects without NAFLD had lower GPR110 mRNA expression than healthy lean subjects, but obese NAFLD subjects had similar GPR110 mRNA expression level as healthy lean subjects (Figure 9A). Subsequently, by using the same transcriptome dataset, we investigated the correlation in the expression level of GPR110 and SCD1. The expression level of GPR110 was positively correlated with SCD1 in the liver (r = 0.4635, P < 0.05; Figure 9B). To verify the observation, we performed immunohistochemistry staining with liver sections from biopsy-proven patients with mild, moderate, and severe NAFLD, respectively (Table S4). The degree of steatosis was determined by non-alcoholic steatohepatitis clinical research network (NASH CRN) scoring system [34]. Immunostaining analysis demonstrated that hepatic expression of GPR110 protein was higher in the ones with severe steatosis than those with lower degree of NAFLD (Figure 9C). These data collectively suggest that GPR110 expression level correlates to hepatic steatosis in humans as well.

**Figure 9:**
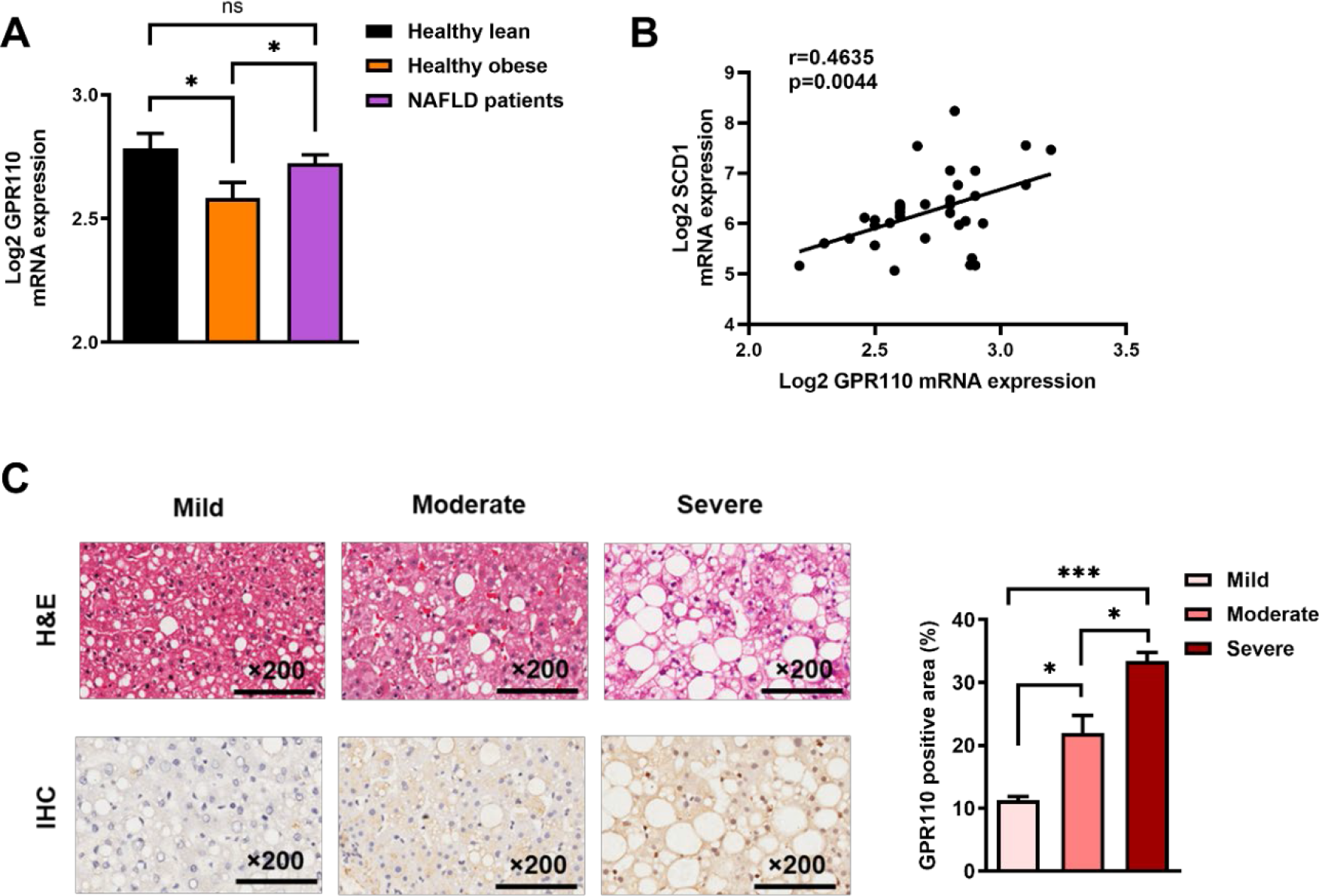
Hepatic expression of GPR110 is upregulated in obese patients with hepatic steatosis when compared to those with normal liver morphology, which is positively associated with hepatic SCD1 expression level. NAFLD patients have higher hepatic expression of GPR110 accompanied with increased mRNA SCD1 expression. (A) Normalized Log2 mRNA expression of GPR110 in lean people without NAFLD (n = 12), obese people without NAFLD (n = 17) or obese patients with NAFLD (n = 8) according to the GEO database (GEO; Profile # GDS4881 / 8126820). (B) Correlation between GPR110 and SCD1 in liver of human subjects based on the GEO database. (C) Representative images of liver tissues with H&E staining (upper panels) and immunohistochemical staining (IHC) of GPR110 (lower panels) from patients with different degree of NAFLD (200 µm). The percentage of GPR110 positive area according to H&E staining (right panel). The percentage of GPR110 positive areas according to IHC staining (right panel); n = 3 per group. Data represents as mean ± SEM. P value analyzed by two-tailed Student’s t test. *P < 0.05, **P < 0.01, ***P < 0.001.

## 4. Discussion

The present study has uncovered a previously unrecognized role of GPR110 in regulating hepatic lipid metabolism. Firstly, we demonstrated that GPR110 is required for regulating lipid content in liver of diet-induced obese mice by both gain-of-function and loss-of-function approaches. HFD-induced steatosis and liver injury were exacerbated in obese mice with high GPR110 overexpression level, and knockdown hepatic GPR110 alleviated the severity of obesity-induced NAFLD. We also confirmed a correlation between hepatic GPR110 expression level and liver steatosis in human. We believe the downregulation of hepatic GPR110 expression level in obese subjects is a protective mechanism to prevent over-accumulation of lipid in liver. It remains to be explored how the transcription level of hepatic GPR110 repressed is in the health obese subjects. In addition, although NAFLD is usually associated with obesity, up to 19% of lean Asian can also present with NAFLD [44]. It is interesting to further explore whether the expression levels of hepatic GPR110 mRNA in these “lean NAFLD” patients are higher than the lean healthy controls.

Subsequently, we also revealed the mechanism which at least in part achieved through the expression of SCD1 in liver. We performed RNA-sequencing analysis to decipher the mechanism and found the expression levels SCD1 mRNAs and protein are dramatically upregulated in the livers of rAAV-GPR110 mice and repressed in GPR110-ASOs treated rAAV-GPR110 mice. SCD1 is the rate- limiting enzyme for catalysing the conversion of saturated long-chain fatty acids into monounsaturated fatty acids. To illustrate that the changes in metabolic phenotype in the rAAV-GPR110 mice were caused by the up-regulation of hepatic SCD1 expression levels, we used SCD1 shRNAs and inhibitor to check whether the metabolic changes can be rescued in GPR110 overexpressing hepatocytes by both *in vivo* and *in vitro* experiments. Concordantly, pharmacologically inhibiting SCD1 by MK8245 was sufficient to rescue the key metabolic dysregulations by overexpressing GPR110 in their livers. Therefore, we concluded that GPR110 induces SCD1 expression, leading to the increase level of *de novo* lipogenesis in liver and exacerbating obese-induced NAFLD.

Previous studies reported that SCD1 global knockout (KO) mice showed improved insulin sensitivity, higher-energy metabolism, and more resistant to diet-induced obesity by the activation of lipid oxidation in addition to the reduction of triglyceride synthesis and storage [45–47]. In addition, the *ob/ob* mice with SCD1 mutations had significantly reduced storage of triglyceride and lower level of very low-density lipoprotein (VLDL) production [48]. Remarkably, liver-specific KO of SCD1 was sufficient to reduce high-carbohydrate diet-induced adiposity with a significant reduction of hepatic lipogenesis and improved glucose tolerance [46]. Indeed, SCD1 inhibition was proposed to be a therapeutic strategy for the treatment of metabolic syndrome [49].

What are the potential advantages of targeting GRP110 rather than SCD1 for the treatment of NAFLD? First of all, SCD1 is highly expressed in various tissues, especially adipose tissues [50]. In addition, expression level and activity of SCD1 is very tightly regulated [51, 52]. Harmful consequences from inhibiting SCD1 have been reported, such as the inhibition of fat mobilization in adipose tissues, and the promotion of proinflammatory and endoplasmic reticulum stress by accumulation SCD1 substrates [53–56]. These findings clearly documented that optimal level of SCD1 is required to maintain health. Secondly, in contrast to SCD1, according to the phenotypes of GPR110 KO mice in previous studies [20, 22] and a dramatical reduction of GPR110 in the livers of HFD-fed mice and health obese subjects, the hepatic GPR110 may be dispensable in adults. Therefore, targeting hepatic GPR110 is a potential safe treatment of NALFD. Thirdly, according to our RNA-sequencing analysis, repressing GPR110 can also regulate the expression of many other lipid metabolism genes. Unfortunately, GPR110 antagonist is not available at this moment. As demonstrated in this study, the ASO-based strategy is an alternative approach to knockdown the expression of GPR110 in liver [57].

The current study focuses on GPR110 in hepatic lipid metabolism. As mentioned above, GPR110 is primarily understood to be an oncogene [15-21, 58-61]. Notably, recent study reported that deficiency of GPR110 can decelerate carcinogen-induced hepatocarcinogenesis in adult mice [20]. Coincidentally, high expression level of SCD1 is also genetically susceptible to hepatocarcinogenesis [62]. It is highly possible that GPR110 also accelerates carcinogenesis by inducing SCD1 expression level. To prove this hypothesis, we may check the SCD1 expression level in the GPR110 induced cancers.

In summary, we present evidence demonstrating a novel role of hepatic GPR110 in regulating lipid metabolism and explored the mechanism partially via regulation of SCD1 expression level. As the amino acid sequencing of GPR110 are highly conserved in humans and mice, targeting GPR110 may serve as a novel therapeutic approach for the treatment of NAFLD patients.

## Data Availability

All data generated or analysed during this study are included in the manuscript and supporting file as Supplementary Table 3.

## Acknowledgement

This work was supported by National Natural Science Foundation of China (81870586), Area of Excellence (AoE/M-707/18), and General Research Fund (15101520) to C.M.W., and National Natural Science Foundation of China (82270941, 81974117) to S.J. We also thank Dr. Oscar Wong and Prof. Zou Xiang for critical review of this manuscript.

## CRediT authorship contribution statement

M.W., A.X. and C.M.W. designed the experiments; M.W., T.H.L., C.D., K.Y.C., X.L. and J.L. conducted the experiments and analysed the data; M.W. and C.M.W. drafted the manuscript; L.L., S.J., S.Y. and P.L. provided essential experimental materials; C.M.W. supervised the whole project.

---

**Supplementary Figure 1:**
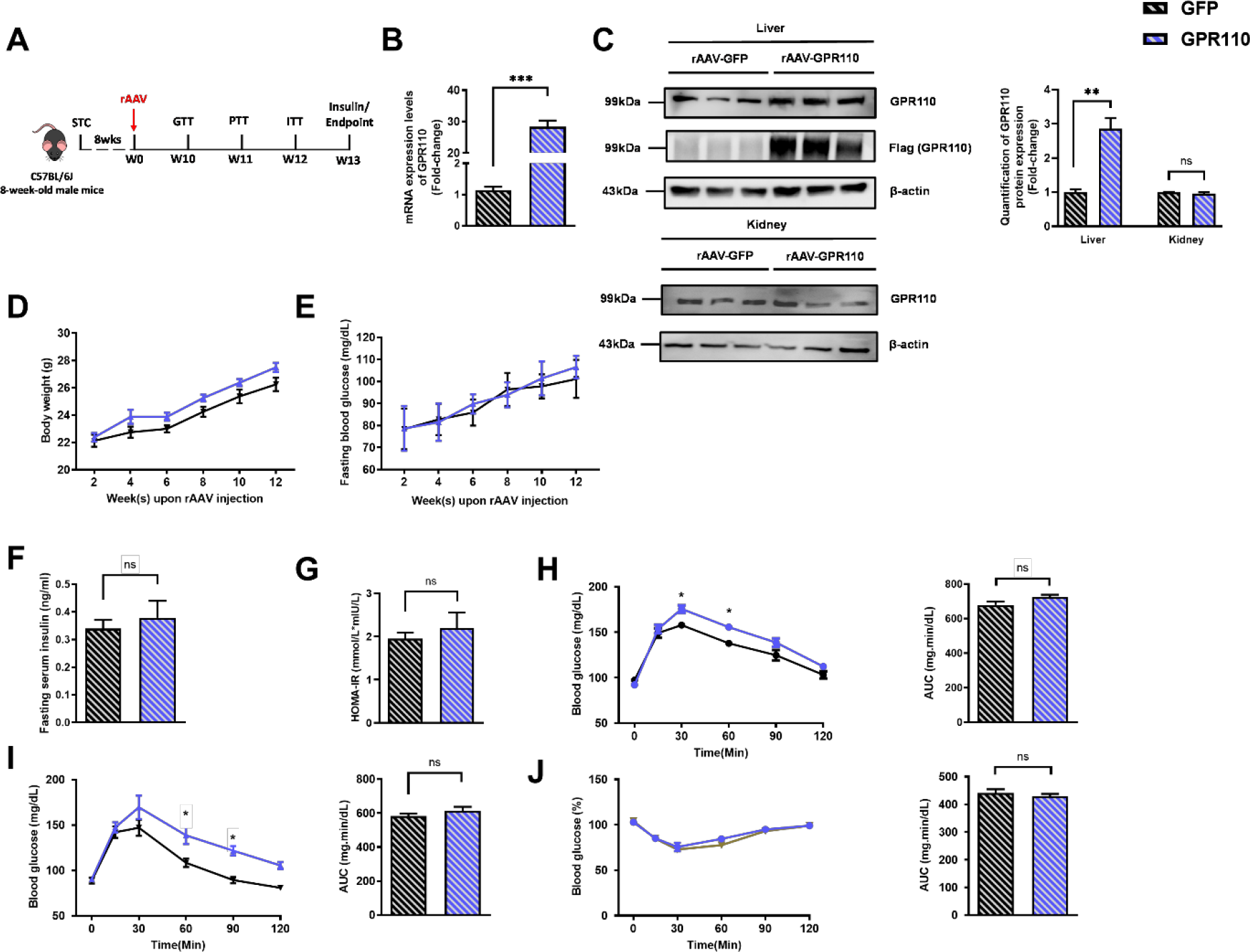
Related to Fig. 2. Hepatic overexpression of GPR110 in STC-fed mice exhibits mild metabolic abnormalities. Eight-week-old male C57BL/6J mice were infected with 3×10^11^ copies of AAV encoding GPR110 (rAAV-GPR110, i.v.) or control (rAAV-GFP, i.v.) and fed with STC diet. (A) Schematic illustration of viral treatments. (B) Hepatic mRNA expression levels of GPR110 from STC- fed mice with liver-specific GPR110 overexpression as determined by qPCR analysis. (C) Left panel: immunoblotting analysis of hepatic protein expression level of GPR110 from STC-fed mice liver with GPR110 overexpression. Right panel: quantification of hepatic protein expression levels of GPR110. Each lane is a sample from a different individual; n = 3 per group. (D) Change of body weight and (E) fasting blood glucose at different weeks upon rAAV injection were measured. (F) Fasting blood insulin level and (G) HOMA-IR values were measured and calculated at the end of the experiment. (H) GTT (1 g/kg BW, left) and area under curve (AUC, right) of serum glucose at the week of 10. (I) PTT (1 g/kg BW, left) and AUC (right) of serum glucose at week 11. (J) ITT (0.5 U/kg BW, left) and AUC (right) of serum glucose at week 12. mRNA expression levels of the target genes were normalized to the expression of mouse GAPDH. STC, standard chow diet; BW, body weight; i.v., intravenous injection; GTT, glucose tolerance test; PTT, pyruvate tolerance test; ITT, insulin tolerance test; AUC, area under curve; NC, negative control; HOMA-IR, homeostasis model assessment-estimated insulin resistance. Data represents as mean ± SEM; n = 8 mice per group; repeated with three independent experiments; P value analyzed by two-tailed Student’s t test. *P < 0.05, **P < 0.01, ***P < 0.001.

**Supplementary Figure 2:**
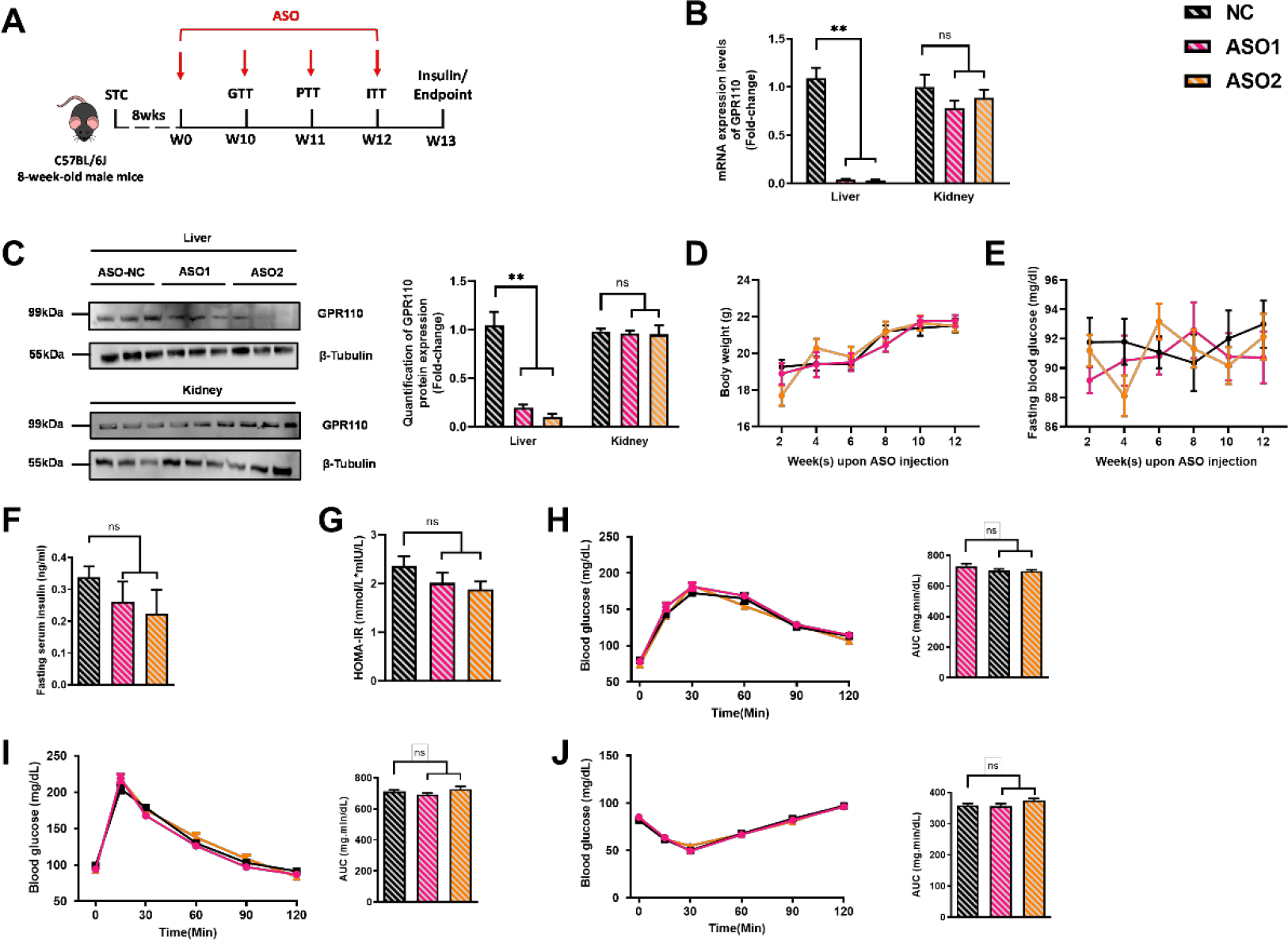
Related to Figure 3. Hepatic knockdown of GPR110 in STC-fed mice does not exhibit metabolic abnormalities. Eight-week-old male C57BL/6J mice were infected with two different sequences of GPR110 antisense oligonucleotides (ASO1-GPR110, ASO2-GPR110, 5 mg/kg BW, one dose per week, s.c.) or scrambled control (ASO-NC, s.c.) received STC feeding, respectively. (A) Schematic illustration of viral treatments. (B) mRNA expression levels of GPR110 in liver and kidney as determined by qPCR analysis. mRNA expression levels of GPR110 in different tissues were normalized to the expression of mouse GAPDH. (C) Left panel: immunoblotting analysis of hepatic protein expression level of GPR110 from STC-fed mice liver with GPR110 knockdown. Right panel: quantification of hepatic protein expression levels of GPR110. Each lane is a sample from a different individual; n = 3 per group. (D) Change of body weight and (E) fasting blood glucose at different weeks upon rAAV injection were measured. (F) Fasting blood insulin level and (G) HOMA-IR values were measured and calculated at the end of the experiment. (H) GTT (1 g/kg BW, left) and area under curve (AUC, right) of serum glucose at the week of 10. (I) PTT (1 g/kg BW, left) and AUC (right) of serum glucose at week 11. (J) ITT (0.5 U/kg BW, left) and AUC (right) of serum glucose at week of 12. mRNA expression levels of the target genes were normalized to the expression of mouse GAPDH. STC, standard chow diet; s.c., subcutaneous injection; ASO, antisense oligonucleotides; BW, body weight; GTT, glucose tolerance test; PTT, pyruvate tolerance test; ITT, insulin tolerance test; AUC, area under curve; NC, negative control; HOMA-IR, homeostasis model assessment- estimated insulin resistance. Data represents as mean ± SEM; n = 8 mice per group; repeated with three independent experiments; P value analyzed by two-tailed Student’s t test. *P < 0.05, **P < 0.01, ***P < 0.001.

**Table S1.**
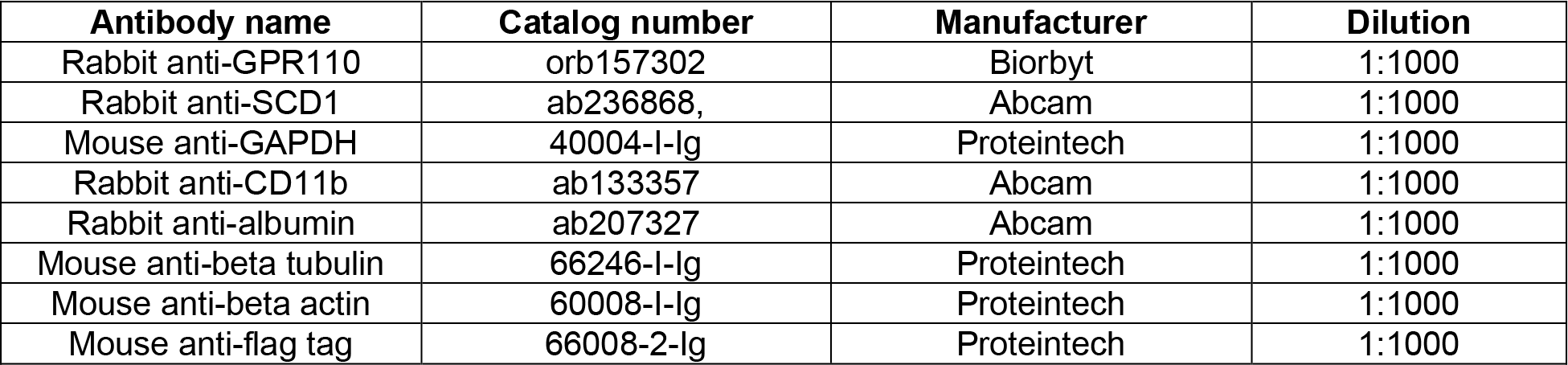
List of primary antibodies

| **Antibody name** | **Catalog number** | **Manufacturer** | **Dilution** |
| --- | --- | --- | --- |
| Rabbit anti-GPR110 | orb157302 | Biorbyt | 1:1000 |
| Rabbit anti-SCD1 | ab236868, | Abcam | 1:1000 |
| Mouse anti-GAPDH | 40004-I-Ig | Proteintech | 1:1000 |
| Rabbit anti-CD11b | ab133357 | Abcam | 1:1000 |
| Rabbit anti-albumin | ab207327 | Abcam | 1:1000 |
| Mouse anti-beta tubulin | 66246-I-Ig | Proteintech | 1:1000 |
| Mouse anti-beta actin | 60008-I-Ig | Proteintech | 1:1000 |
| Mouse anti-flag tag | 66008-2-Ig | Proteintech | 1:1000 |

**Table S2.**
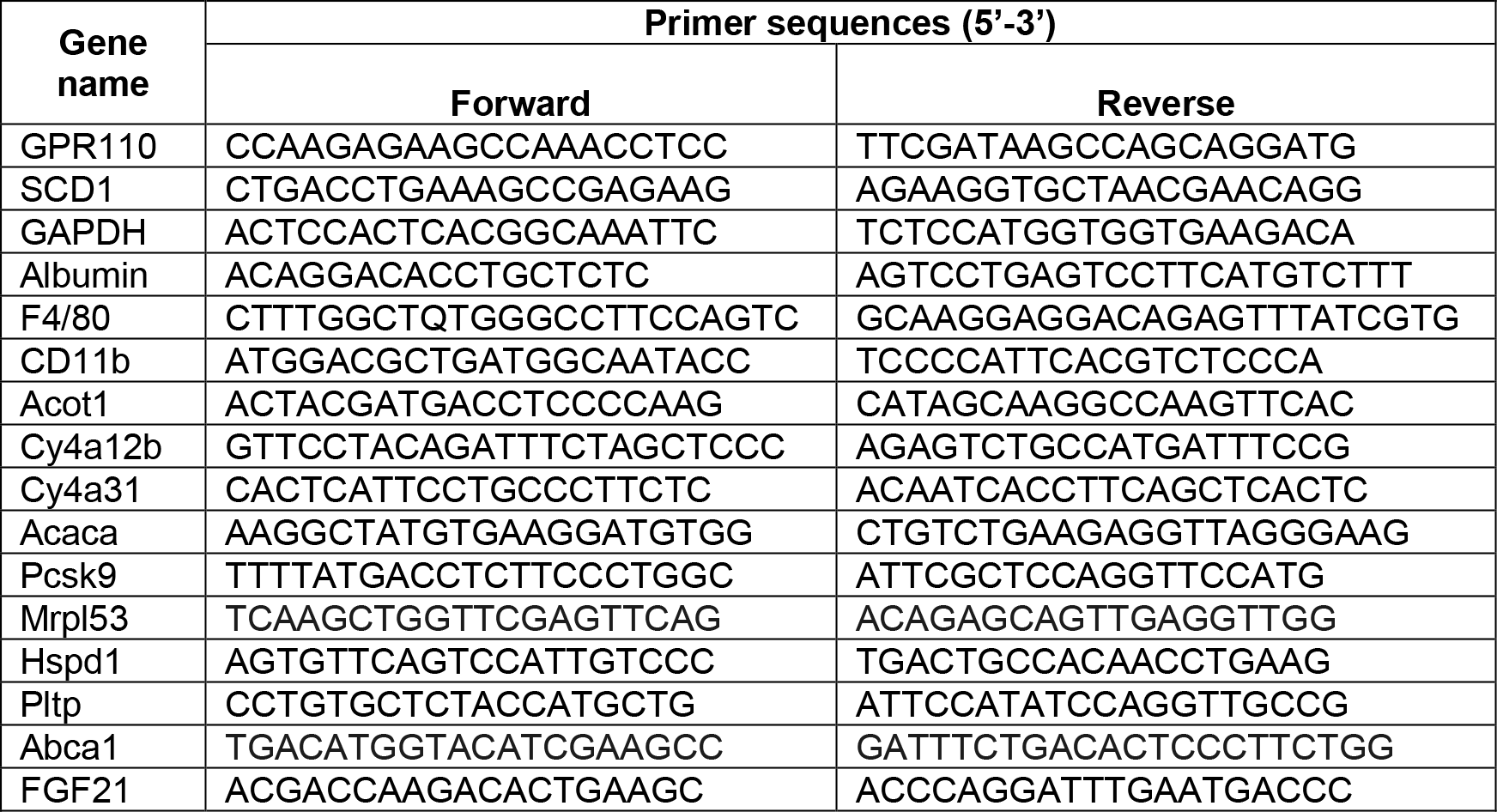
List of primers used for qPCR

**Supplementary Table 3:**
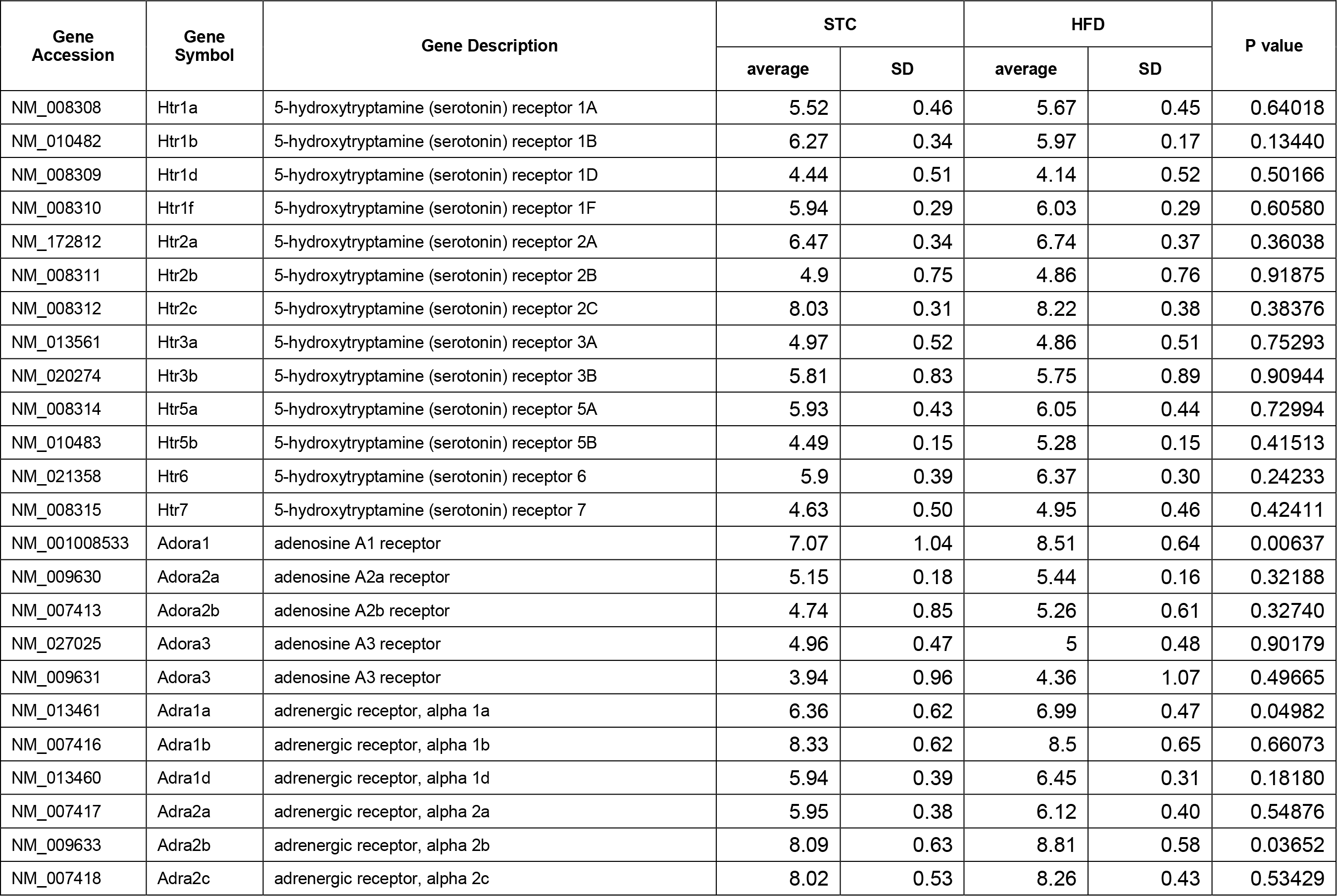
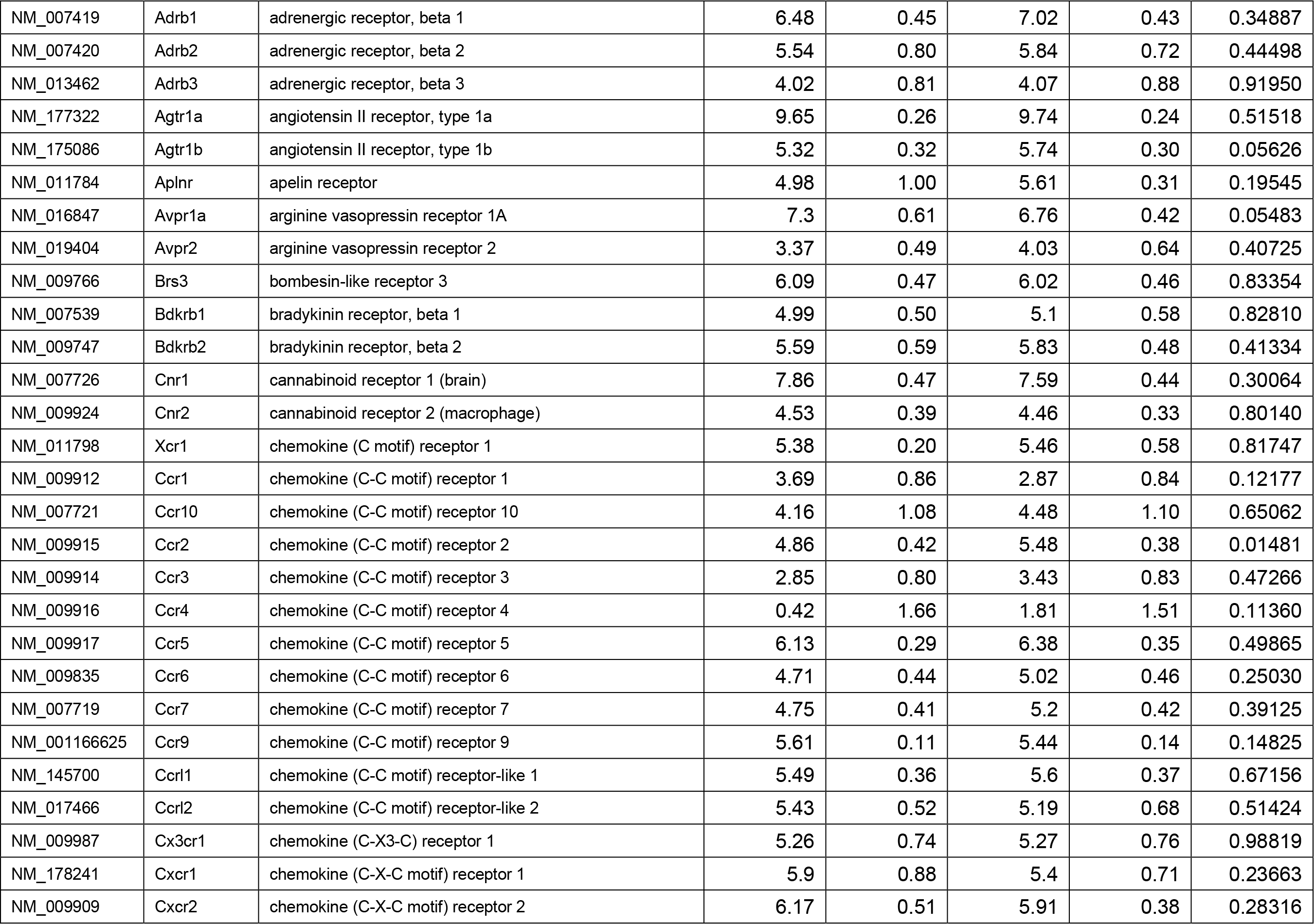
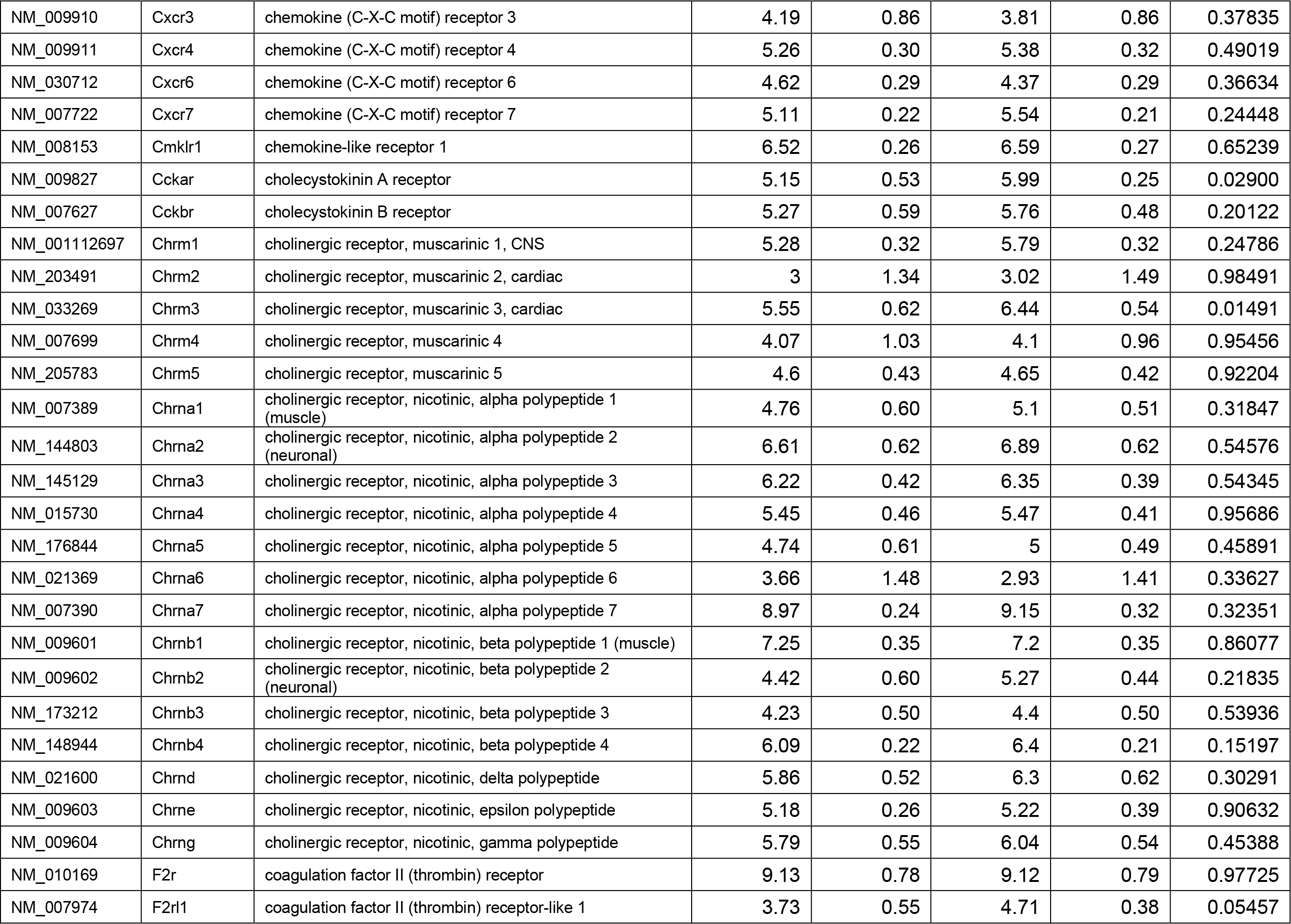
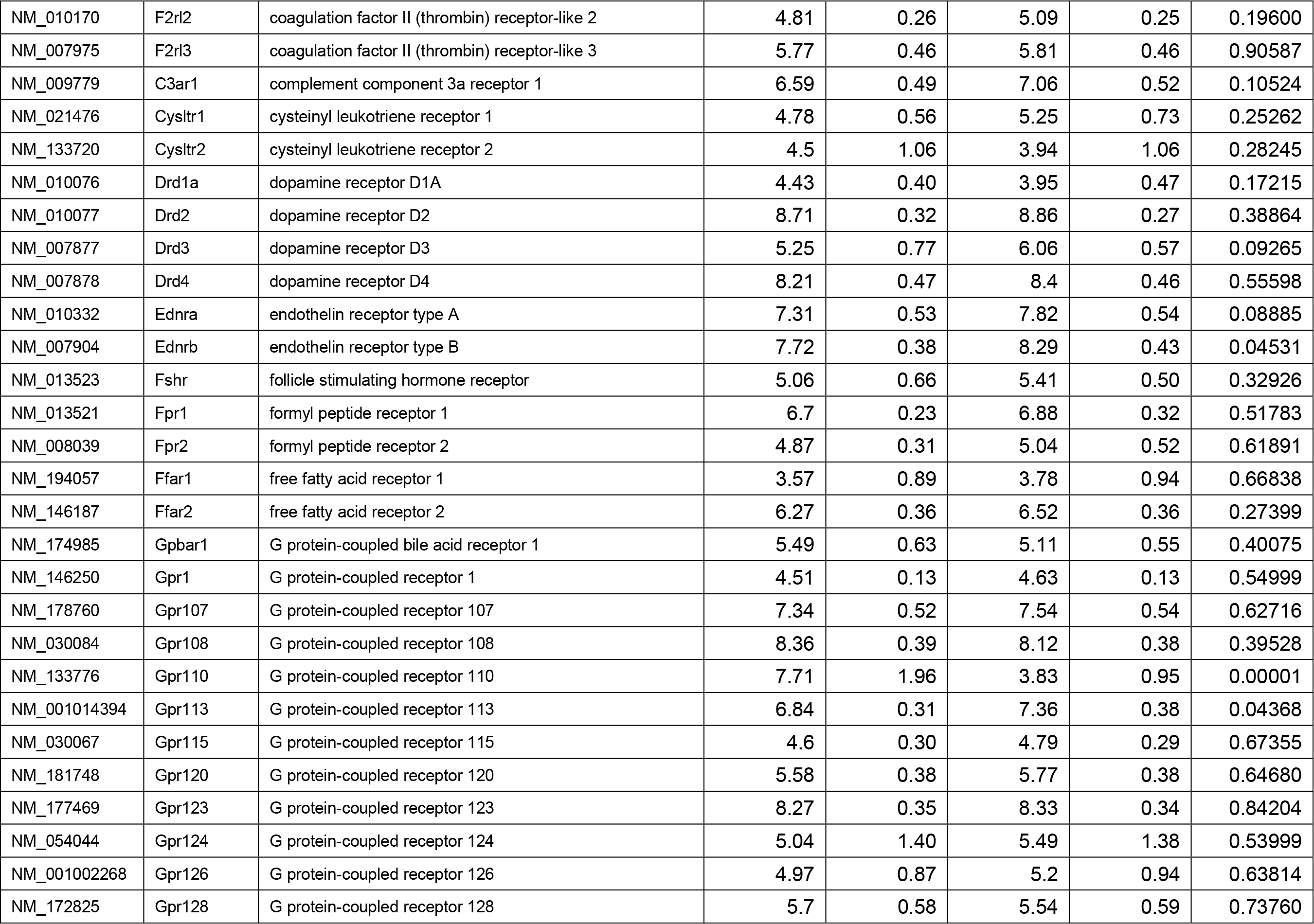
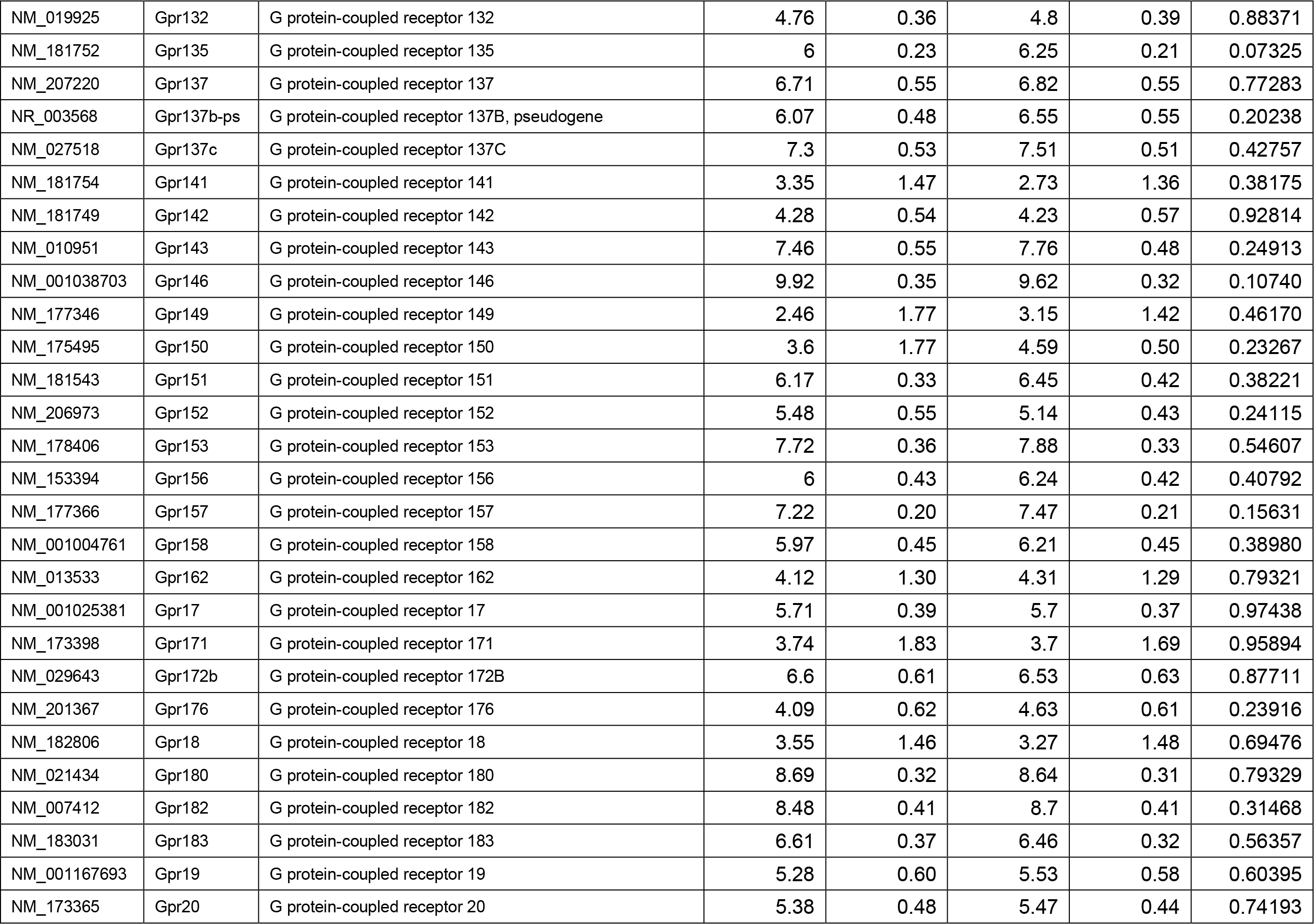
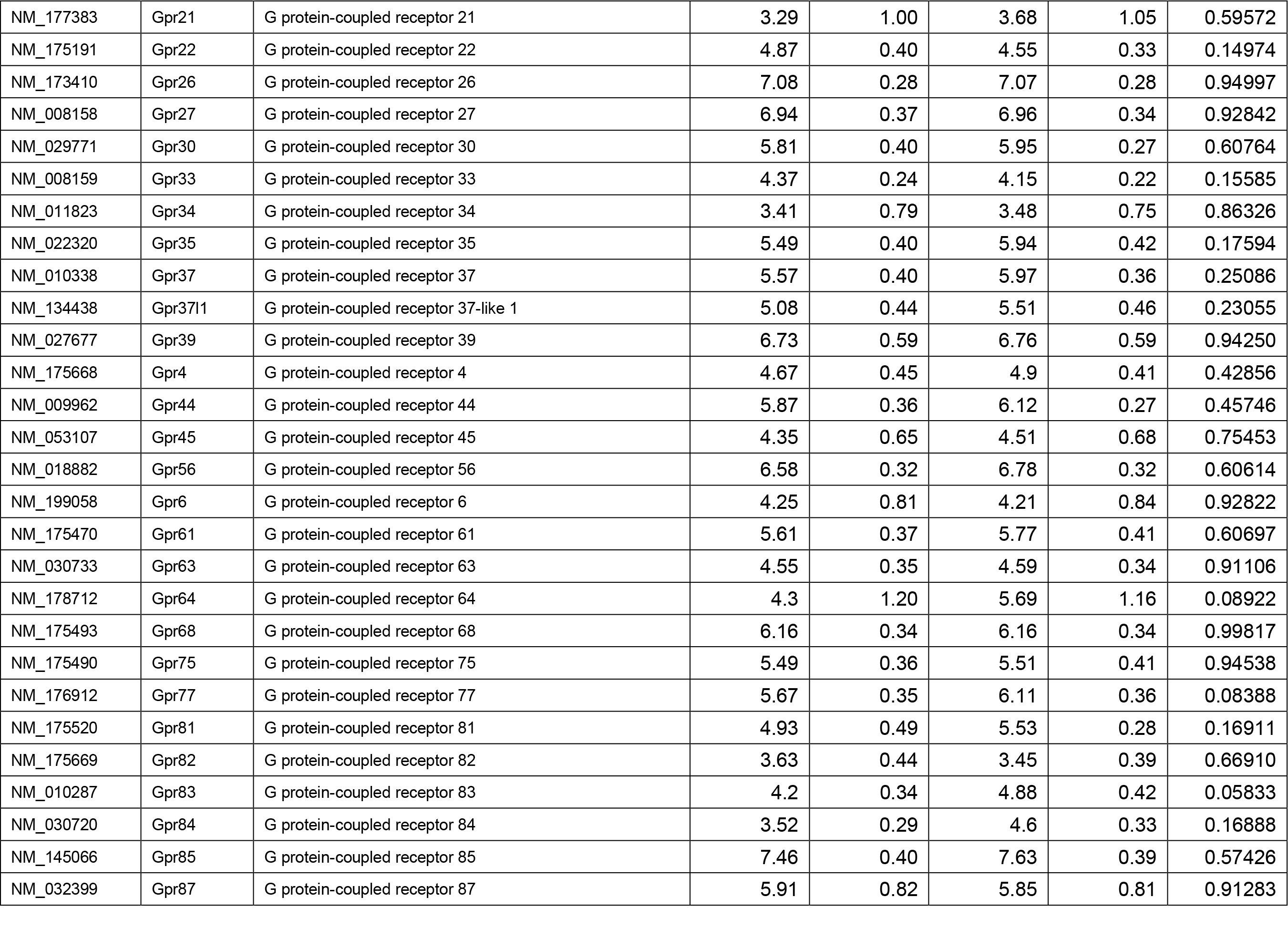
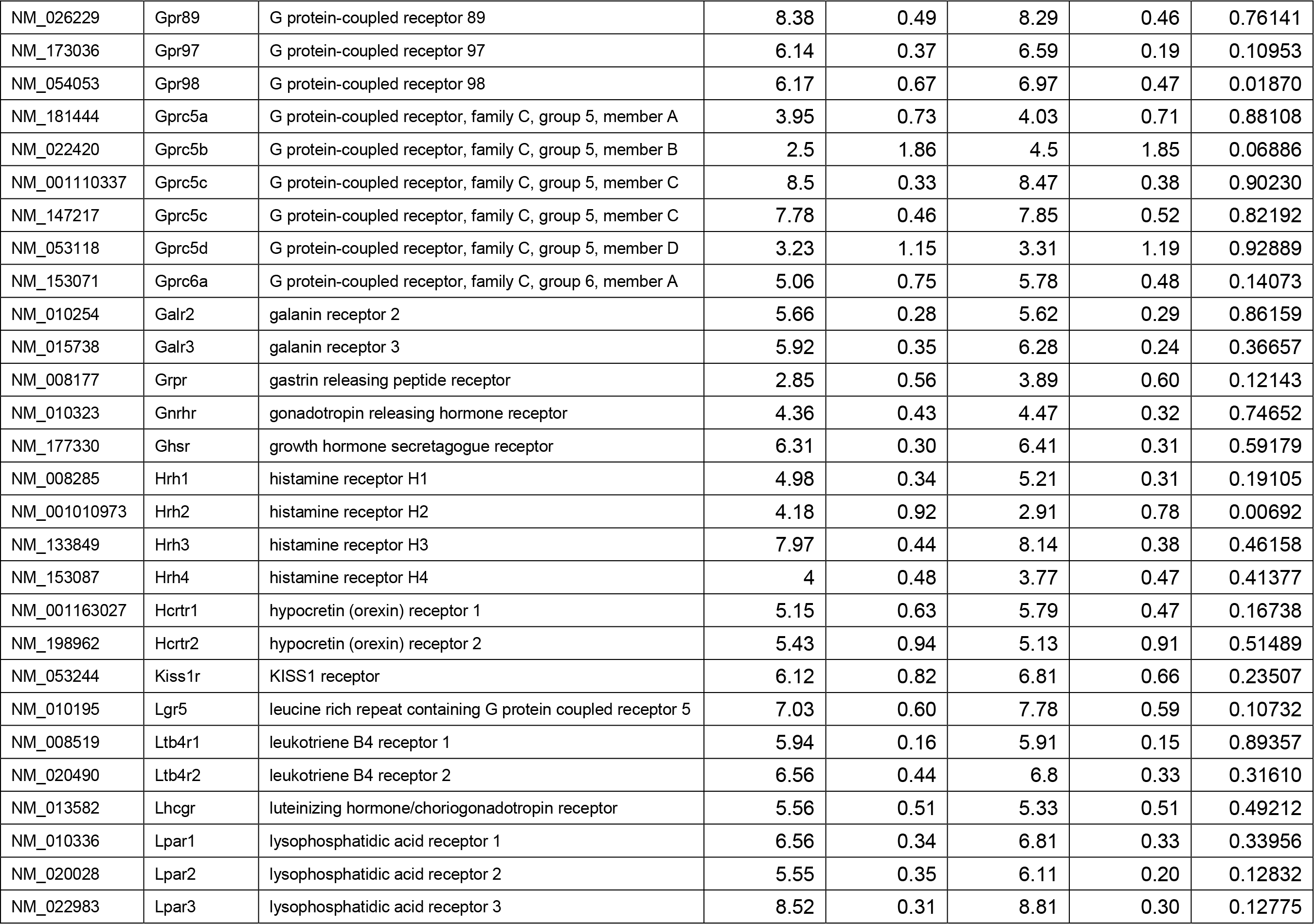
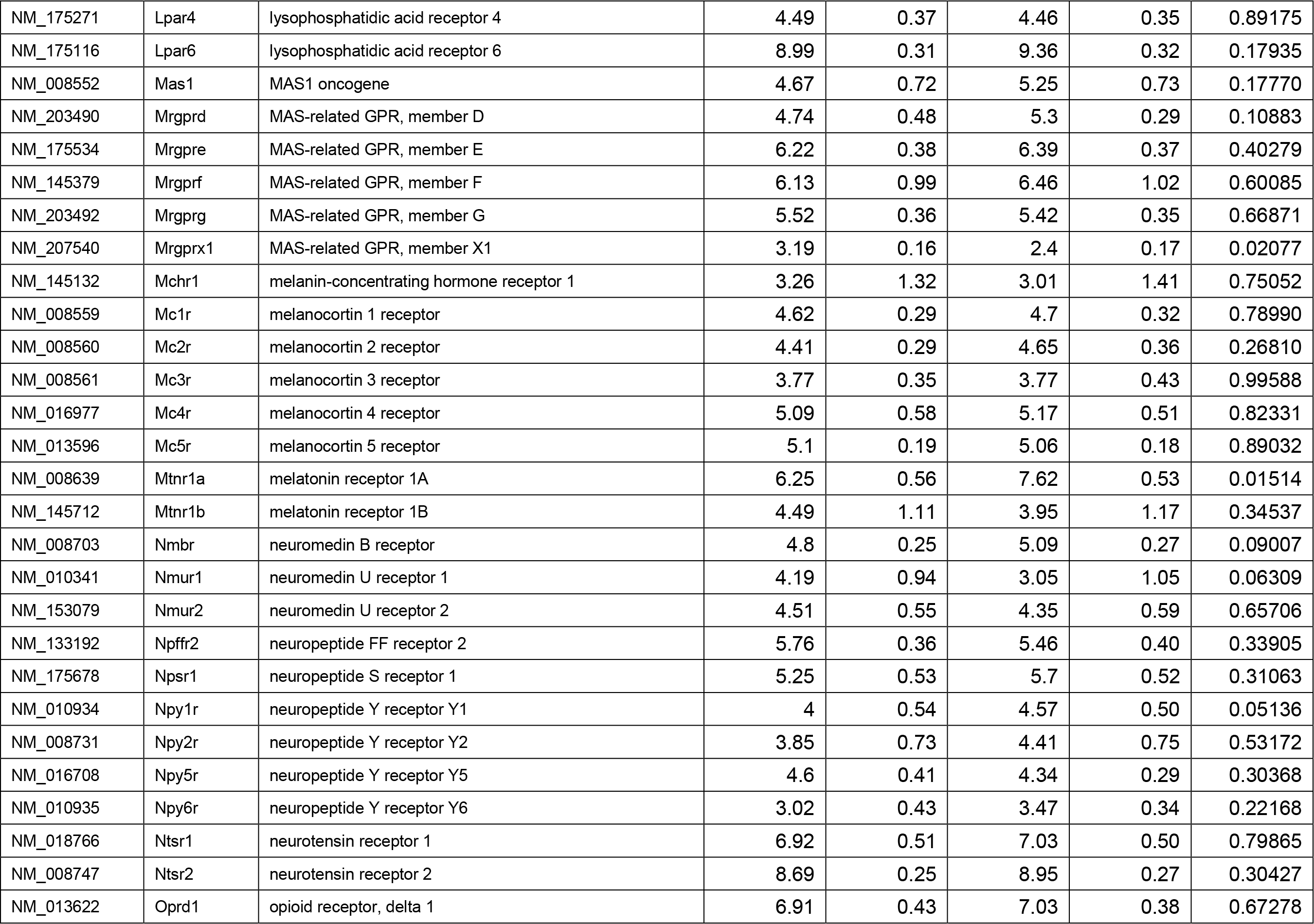
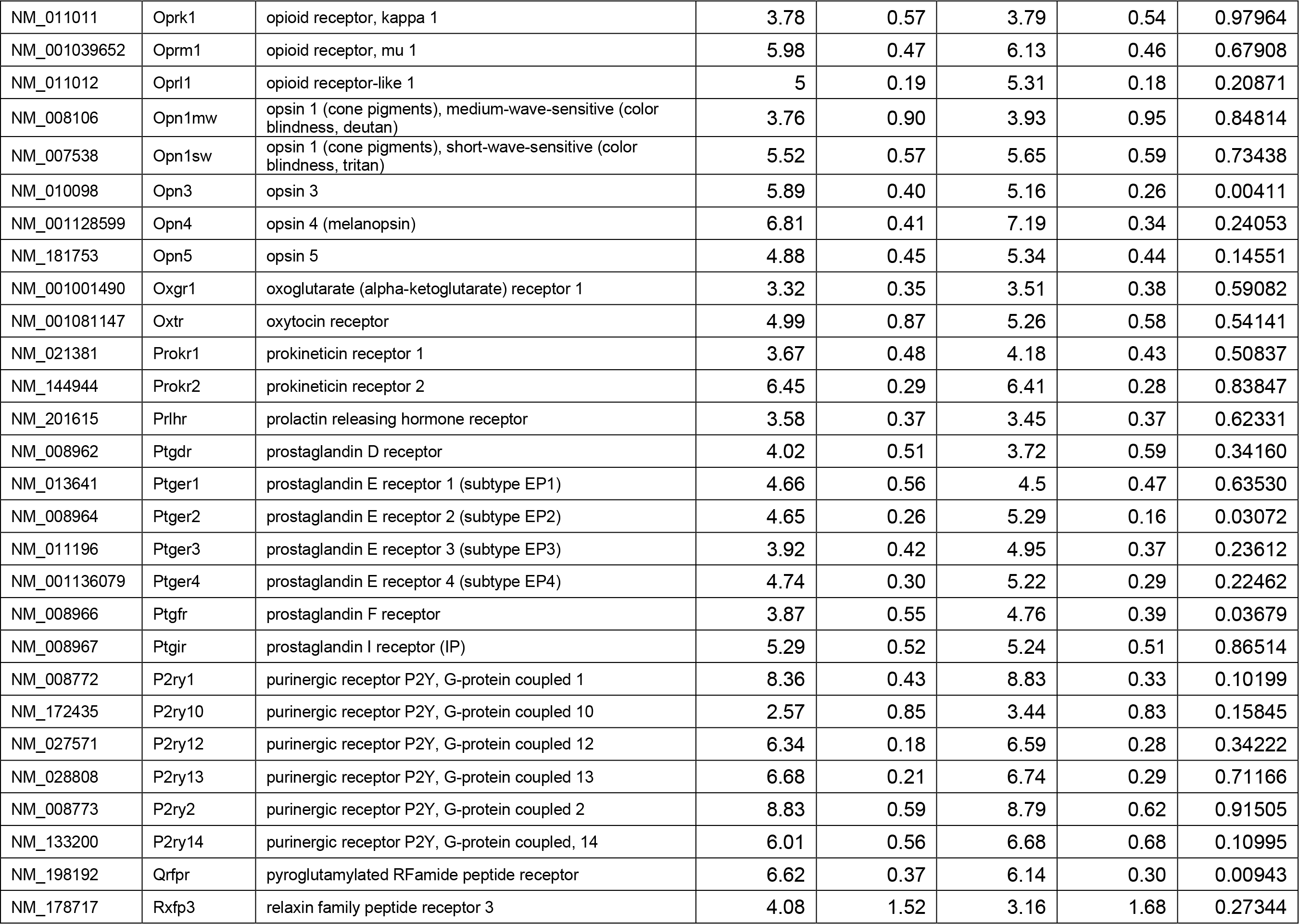
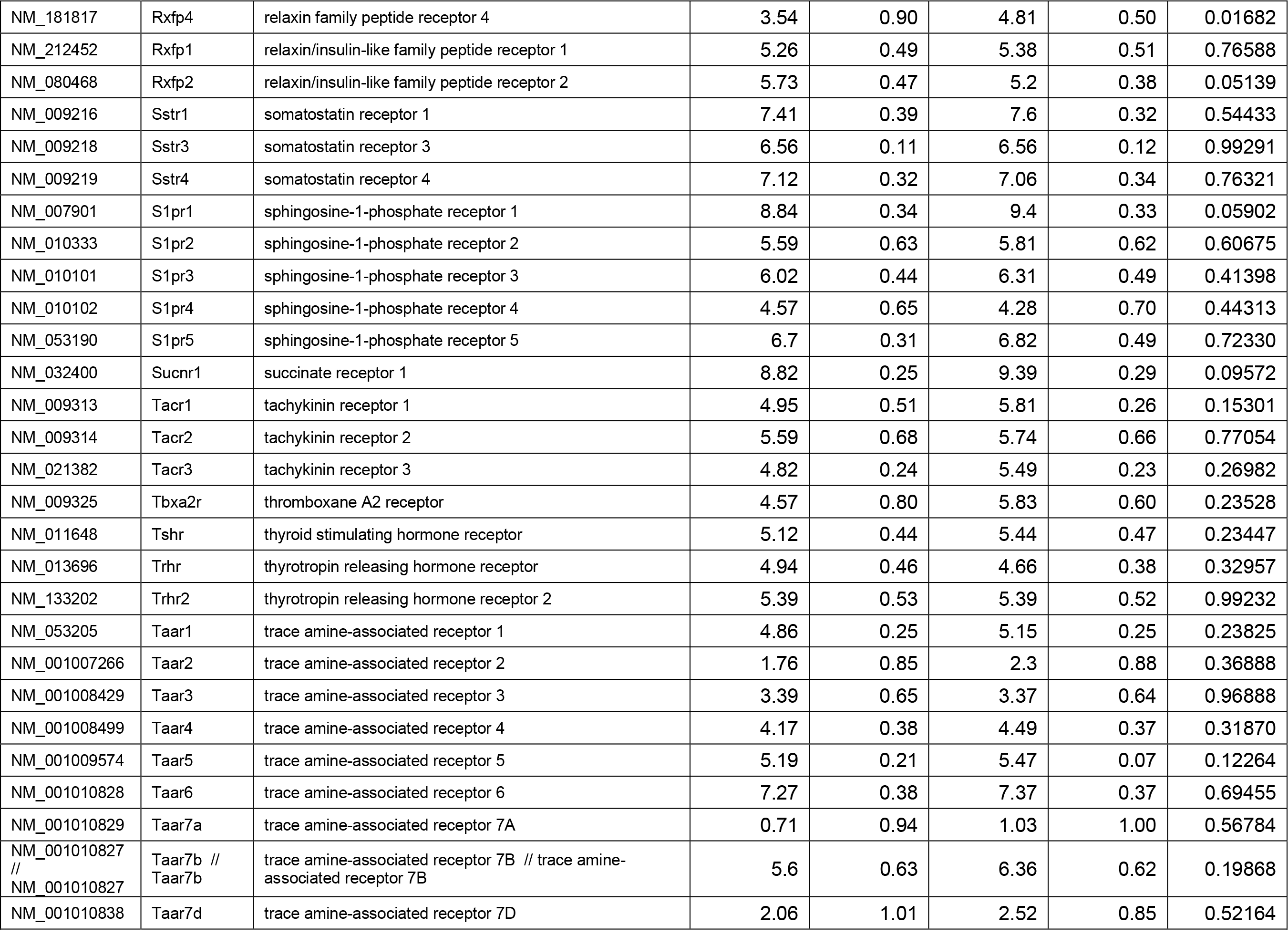
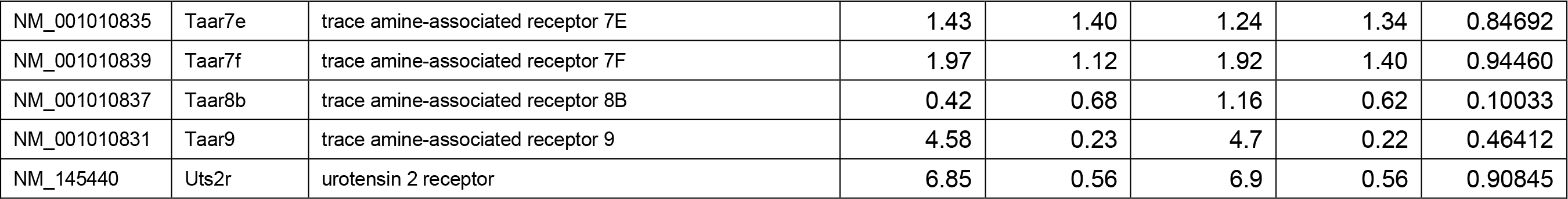
Expression of GPCRs in the liver of mice fed with either STC or HFD diet for 8 weeks by gene expression microarray analysis (Affymetrix Mouse Exon 1.0 ST Array).

**Supplementary Table 4:** Baseline characteristics of study cohorts.

| **NAFLD stage** | **Mild** | **Moderate** | **Severe** |
| --- | --- | --- | --- |
| **Age** | 49.00 ± 15.10 | 47.33 ± 15.89 | 25.67 ± 5.66 |
| **Gender (Female/Male)** | 2/1 | 0/3 | 2/1 |
| **Body Weight (kg)** | 66.67 ± 14.41 | 65.90 ± 15.79 | 79.00 ± 1.41 |
| **Body Height (m)** | 1.60 ± 0.13 | 1.70 ± 0.10 | 1.68 ± 0.01 |
| **BMI (kg/m<sup>2</sup>)** | 25.65 ± 2.06 | 22.59 ± 3.84 | 28.08 ± 0.27 |
| **Fasting Glucose (mmol/L)** | 5.07 ± 0.35 | 6.42 ± 1.42 | 5.51 ± 1.48 |
| **SBP (mmHg)** | 122.33 ± 11.24 | 114.33 ± 26.50 | 130.67 ± 0.71 |
| **DPB (mmHg)** | 76.33 ± 17.04 | 75.33 ± 17.01 | 94.00 ± 25.46 |
| **Total Cholesterol (mmol/L)** | 4.29 ± 0.90 | 3.32 ± 0.52 | 3.93 ± 1.18 |
| **Triglycerides (mmol/L)** | 1.29 ± 0.47 | 1.99 ± 1.44 | 1.06 ± 0.26 |
| **HDL (mmol/L)** | 1.38 ± 0.23 <sup>a</sup> | 0.82 ± 0.25 | 1.05 ± 0.14 |
| **LDL (mmol/L)** | 2.39 ± 0.68 | 1.82 ± 0.21 | 2.51 ± 1.28 |
| **ALT (U/L)** | 38.33 ± 27.39 | 87.33 ± 41.53 | 91.33 ± 52.33 |
| **AST (U/L)** | 16.33 ± 6.81 <sup>a,b</sup> | 46.00 ± 16.00 | 51.33 ± 16.26 |
| **AST/ALT** | 0.63 ± 0.40 | 0.63 ± 0.40 | 0.60 ± 0.21 |
| **γGGT (U/L)** | 117.67 ± 163.10 | 47.67 ± 6.43 | 100.67 ± 32.53 |

Abbreviations: BMI, body mass index; HOMA-IR, homeostasis model assessment of insulin resistance; SBP, systolic blood pressure; DBP, diastolic blood pressure; HDL, high density lipoproteins; LDL, low density lipoproteins; ALT, alanine aminotransferase; AST, aspartate transaminase; γGGT, γ-glutamyl transpeptidase. Data represent as mean ± SEM; a, significant difference between mild and moderate; b, significant difference between mild and severe, no significant differences among the rest groups. P < 0.05 is considered significant.

## References

1. Younossi ZM, Koenig AB, Abdelatif D, Fazel Y, Henry L, Wymer M. Global epidemiology of nonalcoholic fatty liver disease-Meta-analytic assessment of prevalence, incidence, and outcomes. Hepatology 2016;64:73–84.

2. Cotter TG, Rinella M. Nonalcoholic Fatty Liver Disease 2020: The State of the Disease. Gastroenterology 2020;158:1851–1864.

3. Peng C, Stewart AG, Woodman OL, Ritchie RH, Qin CX. Non-Alcoholic Steatohepatitis: A Review of Its Mechanism, Models and Medical Treatments. Front Pharmacol 2020;11:603926.

4. Allen AM, Hicks SB, Mara KC, Larson JJ, Therneau TM. The Risk of Incident Extrahepatic Cancers is higher in Nonalcoholic Fatty Liver Disease than Obesity - a Longitudinal Cohort Study. J Hepatol 2019.

5. Zhou F, Zhou J, Wang W, Zhang XJ, Ji YX, Zhang P, et al. Unexpected Rapid Increase in the Burden of NAFLD in China From 2008 to 2018: A Systematic Review and Meta-Analysis. Hepatology 2019;70:1119–1133.

6. Rinella ME, Sanyal AJ. Management of NAFLD: a stage-based approach. Nat Rev Gastroenterol Hepatol 2016;13:196–205.

7. Yang D, Zhou Q, Labroska V, Qin S, Darbalaei S, Wu Y, et al. G protein-coupled receptors: structure- and function-based drug discovery. Signal Transduct Target Ther 2021;6:7.

8. Hauser AS, Attwood MM, Rask-Andersen M, Schioth HB, Gloriam DE. Trends in GPCR drug discovery: new agents, targets and indications. Nature reviews Drug discovery 2017;16:829–842.

9. Sriram K, Insel PA. G Protein-Coupled Receptors as Targets for Approved Drugs: How Many Targets and How Many Drugs? Molecular pharmacology 2018;93:251–258.

10. Yang M, Zhang CY. G protein-coupled receptors as potential targets for nonalcoholic fatty liver disease treatment. World J Gastroenterol 2021;27:677–691.

11. Kurtz R, Anderman MF, Shepard BD. GPCRs get fatty: the role of G protein-coupled receptor signaling in the development and progression of nonalcoholic fatty liver disease. American journal of physiology Gastrointestinal and liver physiology 2021;320:G304–G318.

12. Fredriksson R, Lagerstrom MC, Hoglund PJ, Schioth HB. Novel human G protein-coupled receptors with long N-terminals containing GPS domains and Ser/Thr-rich regions. FEBS letters 2002;531:407–414.

13. Bjarnadottir TK, Fredriksson R, Hoglund PJ, Gloriam DE, Lagerstrom MC, Schioth HB. The human and mouse repertoire of the adhesion family of G-protein-coupled receptors. Genomics 2004;84:23–33.

14. Bjarnadottir TK, Geirardsdottir K, Ingemansson M, Mirza MA, Fredriksson R, Schioth HB. Identification of novel splice variants of Adhesion G protein-coupled receptors. Gene 2007;387:38–48.

15. Lum AM, Wang BB, Beck-Engeser GB, Li L, Channa N, Wabl M. Orphan receptor GPR110, an oncogene overexpressed in lung and prostate cancer. BMC cancer 2010;10:40.

16. Liu Z, Zhang G, Zhao C, Li J. Clinical Significance of G Protein-Coupled Receptor 110 (GPR110) as a Novel Prognostic Biomarker in Osteosarcoma. Medical science monitor : international medical journal of experimental and clinical research 2018;24:5216–5224.

17. Zhu X, Huang G, Jin P. Clinicopathological and prognostic significance of aberrant G protein-couple receptor 110 (GPR110) expression in gastric cancer. Pathology, research and practice 2019;215:539–545.

18. Shi H, Zhang S. Expression and prognostic role of orphan receptor GPR110 in glioma. Biochem Biophys Res Commun 2017;491:349–354.

19. Bhat RR, Yadav P, Sahay D, Bhargava DK, Creighton CJ, Yazdanfard S, et al. GPCRs profiling and identification of GPR110 as a potential new target in HER2+ breast cancer. Breast cancer research and treatment 2018;170:279–292.

20. Ma B, Zhu J, Tan J, Mao Y, Tang L, Shen C, et al. Gpr110 deficiency decelerates carcinogen- induced hepatocarcinogenesis via activation of the IL-6/STAT3 pathway. Am J Cancer Res 2017;7:433–447.

21. Nam HJ, Kim YJ, Kang JH, Lee SJ. GPR110 promotes progression and metastasis of triple- negative breast cancer. Cell Death Discov 2022;8:271.

22. Lee JW, Huang BX, Kwon H, Rashid MA, Kharebava G, Desai A, et al. Orphan GPR110 (ADGRF1) targeted by N-docosahexaenoylethanolamine in development of neurons and cognitive function. Nature communications 2016;7:13123.

23. Nakamura A, Terauchi Y. Lessons from mouse models of high-fat diet-induced NAFLD. International journal of molecular sciences 2013;14:21240–21257.

24. Sellmann C, Baumann A, Brandt A, Jin CJ, Nier A, Bergheim I. Oral Supplementation of Glutamine Attenuates the Progression of Nonalcoholic Steatohepatitis in C57BL/6J Mice. The Journal of nutrition 2017;147:2041–2049.

25. Lee JT, Huang Z, Pan K, Zhang HJ, Woo CW, Xu A, et al. Adipose-derived lipocalin 14 alleviates hyperglycaemia by suppressing both adipocyte glycerol efflux and hepatic gluconeogenesis in mice. Diabetologia 2016;59:604–613.

26. Cheng Y, Kang XZ, Cheng T, Ye ZW, Tipoe GL, Yu CH, et al. FACI Is a Novel CREB-H-Induced Protein That Inhibits Intestinal Lipid Absorption and Reverses Diet-Induced Obesity. Cell Mol Gastroenterol Hepatol 2022;13:1365–1391.

27. Iida T, Ubukata M, Mitani I, Nakagawa Y, Maeda K, Imai H, et al. Discovery of potent liver-selective stearoyl-CoA desaturase-1 (SCD1) inhibitors, thiazole-4-acetic acid derivatives, for the treatment of diabetes, hepatic steatosis, and obesity. European journal of medicinal chemistry 2018;158:832–852.

28. Xu L, Huang Z, Lo T-h, Lee JTH, Yang R, Yan X, et al. Hepatic PRMT1 ameliorates diet-induced hepatic steatosis via induction of PGC1α. Theranostics 2022;12:2502.

29. Huang Z, Zhong L, Lee JTH, Zhang J, Wu D, Geng L, et al. The FGF21-CCL11 Axis Mediates Beiging of White Adipose Tissues by Coupling Sympathetic Nervous System to Type 2 Immunity. Cell Metab 2017;26:493–508 e494.

30. Wong CM, Wang Y, Lee JT, Huang Z, Wu D, Xu A, et al. Adropin is a brain membrane-bound protein regulating physical activity via the NB-3/Notch signaling pathway in mice. J Biol Chem 2014;289:25976–25986.

31. Ye D, Li H, Wang Y, Jia W, Zhou J, Fan J, et al. Circulating Fibroblast Growth Factor 21 Is A Sensitive Biomarker for Severe Ischemia/reperfusion Injury in Patients with Liver Transplantation. Sci Rep 2016;6:19776.

32. Chen J, Li J, Yiu JHC, Lam JKW, Wong CM, Dorweiler B, et al. TRIF-dependent Toll-like receptor signaling suppresses Scd1 transcription in hepatocytes and prevents diet-induced hepatic steatosis. Sci Signal 2017;10.

33. Xu L, Huang Z, Lo TH, Lee JTH, Yang R, Yan X, et al. Hepatic PRMT1 ameliorates diet-induced hepatic steatosis via induction of PGC1alpha. Theranostics 2022;12:2502–2518.

34. Brunt EM, Janney CG, Di Bisceglie AM, Neuschwander-Tetri BA, Bacon BR. Nonalcoholic steatohepatitis: a proposal for grading and staging the histological lesions. The American journal of gastroenterology 1999;94:2467–2474.

35. Promel S, Waller-Evans H, Dixon J, Zahn D, Colledge WH, Doran J, et al. Characterization and functional study of a cluster of four highly conserved orphan adhesion-GPCR in mouse. Developmental dynamics : an official publication of the American Association of Anatomists 2012;241:1591–1602.

36. Rusli F, Deelen J, Andriyani E, Boekschoten MV, Lute C, van den Akker EB, et al. Fibroblast growth factor 21 reflects liver fat accumulation and dysregulation of signalling pathways in the liver of C57BL/6J mice. Sci Rep 2016;6:30484.

37. Kopec AK, Abrahams SR, Thornton S, Palumbo JS, Mullins ES, Divanovic S, et al. Thrombin promotes diet-induced obesity through fibrin-driven inflammation. J Clin Invest 2017;127:3152–3166.

38. Cui H, Zhu X, Li S, Wang P, Fang J. Liver-Targeted Delivery of Oligonucleotides with N- Acetylgalactosamine Conjugation. ACS Omega 2021;6:16259–16265.

39. Paton CM, Ntambi JM. Biochemical and physiological function of stearoyl-CoA desaturase. American journal of physiology Endocrinology and metabolism 2009;297:E28–37.

40. Kotronen A, Seppanen-Laakso T, Westerbacka J, Kiviluoto T, Arola J, Ruskeepaa AL, et al. Hepatic stearoyl-CoA desaturase (SCD)-1 activity and diacylglycerol but not ceramide concentrations are increased in the nonalcoholic human fatty liver. Diabetes 2009;58:203–208.

41. Mar-Heyming R, Miyazaki M, Weissglas-Volkov D, Kolaitis NA, Sadaat N, Plaisier C, et al. Association of stearoyl-CoA desaturase 1 activity with familial combined hyperlipidemia. Arterioscler Thromb Vasc Biol 2008;28:1193–1199.

42. Oballa RM, Belair L, Black WC, Bleasby K, Chan CC, Desroches C, et al. Development of a liver- targeted stearoyl-CoA desaturase (SCD) inhibitor (MK-8245) to establish a therapeutic window for the treatment of diabetes and dyslipidemia. Journal of medicinal chemistry 2011;54:5082–5096.

43. Ahrens M, Ammerpohl O, von Schonfels W, Kolarova J, Bens S, Itzel T, et al. DNA methylation analysis in nonalcoholic fatty liver disease suggests distinct disease-specific and remodeling signatures after bariatric surgery. Cell Metab 2013;18:296–302.

44. VanWagner LB, Armstrong MJ. Lean NAFLD: A not so benign condition? Hepatol Commun 2018;2:5–8.

45. Ntambi JM, Miyazaki M, Stoehr JP, Lan H, Kendziorski CM, Yandell BS, et al. Loss of stearoyl- CoA desaturase-1 function protects mice against adiposity. Proc Natl Acad Sci U S A 2002;99:11482–11486.

46. Miyazaki M, Flowers MT, Sampath H, Chu K, Otzelberger C, Liu X, et al. Hepatic stearoyl-CoA desaturase-1 deficiency protects mice from carbohydrate-induced adiposity and hepatic steatosis. Cell Metab 2007;6:484–496.

47. Miyazaki M, Sampath H, Liu X, Flowers MT, Chu K, Dobrzyn A, et al. Stearoyl-CoA desaturase-1 deficiency attenuates obesity and insulin resistance in leptin-resistant obese mice. Biochem Biophys Res Commun 2009;380:818–822.

48. Cohen P, Miyazaki M, Socci ND, Hagge-Greenberg A, Liedtke W, Soukas AA, et al. Role for stearoyl-CoA desaturase-1 in leptin-mediated weight loss. Science 2002;297:240–243.

49. Jiang G, Li Z, Liu F, Ellsworth K, Dallas-Yang Q, Wu M, et al. Prevention of obesity in mice by antisense oligonucleotide inhibitors of stearoyl-CoA desaturase-1. J Clin Invest 2005;115:1030–1038.

50. Ascenzi F, De Vitis C, Maugeri-Sacca M, Napoli C, Ciliberto G, Mancini R. SCD1, autophagy and cancer: implications for therapy. Journal of experimental & clinical cancer research : CR 2021;40:265.

51. AM AL, Syed DN, Ntambi JM. Insights into Stearoyl-CoA Desaturase-1 Regulation of Systemic Metabolism. Trends in endocrinology and metabolism: TEM 2017;28:831–842.

52. Mauvoisin D, Mounier C. Hormonal and nutritional regulation of SCD1 gene expression. Biochimie 2011;93:78–86.

53. Brown JM, Rudel LL. Stearoyl-coenzyme A desaturase 1 inhibition and the metabolic syndrome: considerations for future drug discovery. Current opinion in lipidology 2010;21:192–197.

54. Leung JY, Kim WY. Stearoyl co-A desaturase 1 as a ccRCC therapeutic target: death by stress. Clin Cancer Res 2013;19:3111–3113.

55. Liu X, Strable MS, Ntambi JM. Stearoyl CoA desaturase 1: role in cellular inflammation and stress. Adv Nutr 2011;2:15–22.

56. Zou Y, Wang YN, Ma H, He ZH, Tang Y, Guo L, et al. SCD1 promotes lipid mobilization in subcutaneous white adipose tissue. Journal of lipid research 2020;61:1589–1604.

57. Dhuri K, Bechtold C, Quijano E, Pham H, Gupta A, Vikram A, et al. Antisense Oligonucleotides: An Emerging Area in Drug Discovery and Development. J Clin Med 2020;9.

58. Abdulkareem NM, Bhat R, Qin L, Vasaikar S, Gopinathan A, Mitchell T, et al. A novel role of ADGRF1 (GPR110) in promoting cellular quiescence and chemoresistance in human epidermal growth factor receptor 2-positive breast cancer. The FASEB journal : official publication of the Federation of American Societies for Experimental Biology 2021;35:e21719.

59. Sadras T, Heatley SL, Kok CH, Dang P, Galbraith KM, McClure BJ, et al. Differential expression of MUC4, GPR110 and IL2RA defines two groups of CRLF2-rearranged acute lymphoblastic leukemia patients with distinct secondary lesions. Cancer letters 2017;408:92–101.

60. Espinal-Enriquez J, Munoz-Montero S, Imaz-Rosshandler I, Huerta-Verde A, Mejia C, Hernandez- Lemus E. Genome-wide expression analysis suggests a crucial role of dysregulation of matrix metalloproteinases pathway in undifferentiated thyroid carcinoma. BMC genomics 2015;16:207.

61. Harvey RC, Mullighan CG, Wang X, Dobbin KK, Davidson GS, Bedrick EJ, et al. Identification of novel cluster groups in pediatric high-risk B-precursor acute lymphoblastic leukemia with gene expression profiling: correlation with genome-wide DNA copy number alterations, clinical characteristics, and outcome. Blood 2010;116:4874–4884.

62. Falvella FS, Pascale RM, Gariboldi M, Manenti G, De Miglio MR, Simile MM, et al. Stearoyl-CoA desaturase 1 (Scd1) gene overexpression is associated with genetic predisposition to hepatocarcinogenesis in mice and rats. Carcinogenesis 2002;23:1933–1936.

